# Penalized Semiparametric Estimation for Causal Inference with Possibly Invalid Instruments

**DOI:** 10.1101/2024.01.19.24301518

**Authors:** Yunlong Cao, Yuquan Wang, Dapeng Shi, Dong Chen, Yue-Qing Hu

## Abstract

Inferring causal effects with unmeasured confounder is a main challenge in causal inference. Many researchers impose parametric assumptions on the distribution of unmeasured confounder. However, due to the unobservable nature of the unmeasured confounder, it is more reasonable to leave its distribution unrestricted. Another key challenge in causal inference is the involvement of invalid instrumental variables, which may lead to biased inference and possibly misleading scientific conclusions. To this end, we employ a flexible semiparametric model that allows for possibly invalid instruments without specifying the distribution of unmeasured confounder in this work. A penalized semiparametric estimator for causal effects is constructed and its oracle and asymptotic properties are well established for statistical inference. We evaluate the performance of the estimator through simulation studies, revealing that our proposed estimator exhibits asymptotic unbiasedness and robustness in estimating causal effects, along with consistent selection of invalid instruments. We also demonstrate its application using Atherosclerosis Risk in Communities Study data set, which further validates its robustness in the presence of invalid instruments. Additionally, we have implemented the proposed method in R, and the corresponding R code is available for free download.

## 1. Introduction

Causal inference is vital for elucidating cause-and-effect relationships. For observational data, it is rather difficult to make inference due to the existence of unmeasured confounders. Instrumental variables (IV) stand out as a widely used technique for detecting causality and estimating the causal effect of an exposure on an outcome in this situation. A valid IV which is suitable for estimating causal effects, must adhere to three fundamental assumptions (Angrist, 1996; Sargan, 1958), namely:

1. Relevance: the IV is related to the exposure;
2. Exchangeability: the IV is independent of unmeasured confounders;
3. Exclusion Restriction: the IV has no direct effect on the outcome.

The relevance of instruments can be scrutinized through observed data of exposure and instruments. However, checking assumptions (2) and (3) in a data-dependent manner necessitates substantial domain expertise to discern valid IVs.

In certain instances, causal effects can be deduced even with the presence of invalid IVs. In the context of a linear outcome model, Kolesár et al. (2015) and Bowden et al. (2015) provided solutions wherein all candidate IVs may be valid, but the strength of the IV and its direct effect on the outcome are nearly orthogonal. Kang et al. (2016) and Windmeijer et al. (2018) put forth consistent estimators for causal effects, assuming the majority rule that at least 50% of the IVs are valid. Li and Guo (2020) extended the majority rule to nonlinear outcome models, presenting the three-step inference procedure SpotIV for estimating the conditional average treatment effect (CATE).

Semiparametric methodologies are extensively employed in causal inference. Sun et al. (2023) introduced a class of g-estimators guaranteed to maintain consistency and asymptotic normality in estimating the causal effect of interest, even in the presence of invalid instrumental variables. Zhang and Tchetgen Tchetgen (2022) proposed a robust estimator reaching the efficiency bound for the semiparametric model, without imposing parametric assumptions on the unmeasured confounder. They established the consistency and asymptotic normality of the estimator under appropriate identification and regularity conditions. However, a general identification condition for the semiparametric model is not explicitly stated.

Considering the assumption of majority rule in semiparametric model setting, the penalized semiparametric estimating approaches were developed to estimate causal effects. Diverging from various penalties imposed on the loss function in the conventional parametric models, the semiparametric approach to estimating causal effects does not involve minimizing any objective function. Fu (2003) proposed penalizing the estimating function, instead of the loss function, for generalized linear models with a bridge penalty (Frank and Friedman, 1993; Fu and Knight, 2000). Subsequently, Johnson et al. (2008) presented a comprehensive asymptotic properties of estimators derived from a broad class of penalized estimating functions.

Given semiparametric model setting and majority rule, we explore the penalized semi-parametric estimating method to simultaneously estimate causal effects and select invalid instrumental variables in this work. The article is organized as follows. Section Methods serves to introduce our model setting and discuss the identifiability of model. Subsequently, we present the semiparametric estimating equations (SEE) for the model and introduce the penalized semiparametric estimating equations (PSEE). Section Implementation and Results establishes the algorithm and asymptotic theory for PSEE. In Section Simulation Study, numerical results from simulation studies are presented. Moving to Section Real Data Analysis, we apply the PSEE method to the Atherosclerosis Risk in Communities Study (ARIC) dataset. Section Discussions is dedicated to providing some discussions.

## 2. Methods

### 2.1. Semiparametric model setting

Consider the causal effect of an exposure *D* ∈ ℝ and outcome *Y* ∈ ℝ. **Z** ∈ ℝ^*q*^ denote the *q*-dimensional vector of instrumental variables for inferring the causality, *U* ∈ ℝ is a scalar unmeasured confounder.

We consider the following outcome model,

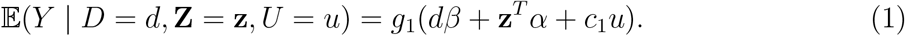

For the exposure *D*, we consider

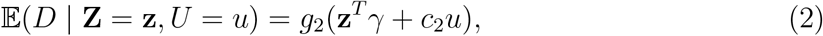

where *g*_1_, *g*_2_ are link functions, *β* ∈ ℝ represents the causal parameter of interest, *α* ∈ ℝ^*q*^ and *γ* ∈ ℝ^*q*^ represent the direct effect of the instruments on the outcome and exposure, respectively, *c*_1_, *c*_2_ are fixed sensitivity parameters used to adjust the influence of confounder on the outcome. Let *θ* = (*γ*^*T*^, *β, α*^*T*^)^*T*^ ∈ ℝ^*p*^ denote the finite dimensional parameters with *p* = 2*q* + 1.

In many applications (Harbord et al., 2013), the distribution of *U* is often assumed to follow a parametric distribution, such as the normal distribution in Shi et al. (2023). Since *U* is unobservable, it may be more appropriate to refrain from imposing any parametric assumptions on its distribution. Therefore we are exploring a semiparametric model in which both the outcome model and exposure model are accurately specified, as denoted by Equations (1) and (2), respectively. The joint distribution of (*U*, **Z**) in this model remains unrestricted.

Note that the model (1)-(2) is highly versatile, encompassing linear and nonlinear outcome as well as exposure as special cases. For example, when *g*_1_ is identity function, it includes continuous outcome as

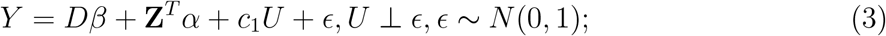

when *g*_1_ is standard logistic function, it includes binary outcome as

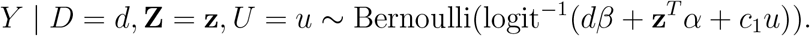

The exposure model (2) is similar, it includes continuous and binary exposure depending on *g*_2_.

### 2.2. Identifiability of Model

The presence of direct effects of instruments on the outcome poses a significant challenge for instrumental variable inference. Previous studies have delved into the identifiability conditions within some models, as discussed by Kang et al. (2016) and Li and Guo (2020). This section provides an overview of these works, laying the foundation for the subsequent discussion of our estimating method for the model parameters.

Let *s* = ∥*α*∥_0_ denote the number of invalid instruments, i.e. the number of nonzero components of *α*. For a continuous outcome, Kang et al. (2016) proved the identifiability of the model under the condition of *s < q/*2, known as the *majority rule*. Additionally, they provided a method sisVIVE for estimating the causal effect, which utilizes *l*_1_ penalization on *α*. Li and Guo (2020) studied the nonlinear causal inference and proposed a method SpotIV to estimate the CATE.

Therefore in this article, we assume that majority rule holds and our model (1)-(2) is identifiable under majority rule. In particular, when outcome *Y* is continuous and Equation (3) holds, and 𝔼(*U*|**Z**) = 0, then our model is actually identifiable under majority rule according to Kang et al. (2016); similarly when exposure *D* is continuous and 𝔼(*U*|**Z**) = 0, our model is identifiable according to Li and Guo (2020).

### 2.3. Semiparametric Estimating Equation

In standard semiparametric theory, we only consider estimators that are regular and asymptotically linear (RAL) (Newey, 1990; Bickel et al., 1993; Van der Vaart, 2000; Tsiatis, 2006). Note that the full data *ℱ* = *{ℱ*_*i*_ = (*Y*_*i*_, *D*_*i*_, **Z**_*i*_, *U*_*i*_), *i* = 1, 2*…, n}*, the observed data only consist of *𝒪* = *{O*_*i*_ = (*Y*_*i*_, *D*_*i*_, **Z**_*i*_), *i* = 1, 2, *…, n}* as *U* is not observed. An asymptotically linear estimator 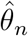 of model parameter *θ* based on the full data satisfies

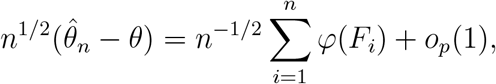

where the measurable random function *φ*(*F*_*i*_) is referred to as the *i*-th influence function of the estimator 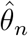 and satisfies *E{φ*(*F*)*}* = 0, *E*(*φφ*^*T*^) is finite and nonsingular. Regularity conditions are imposed to exclude super-efficient estimators, which are unnatural and have undesirable local properties.

Any RAL estimator is asymptotically normally distributed; i.e.,

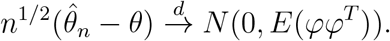

We hope to find the efficient influence function *φ*_eff_ (*F*), which is the influence function with the smallest variance matrix in the sense that for any influence function *φ*(*F*) ≠ *φ*_eff_ (*F*), var{*φ*_eff_ (F)} *−* var{*φ*(F)} is negative definite.

By standard semiparametric theory, the efficient influence function based on the full data, *φ*_eff_ (*F*), is proportional to the full data efficient score *S*_eff_(*Y, D*, **Z**, *U*), which can be obtained by projecting the full data score *S*_*θ*_(*Y, D*, **Z**, *U*) onto the orthogonal component of the full data nuisance tangent space Λ^*F*^, which is given by 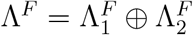, where

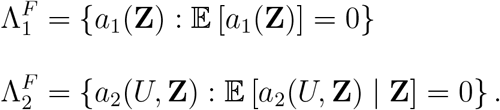

Accordingly, for observed data efficient score *S*_eff_(*Y, D*, **Z**), we need to calculate corresponding observed data score *S*_*θ*_(*Y, D*, **Z**) = 𝔼[*S*_*θ*_(*Y, D*, **Z**, *U*) | *Y, D*, **Z**] and the observed data nuisance tangent space Λ = 𝔼[Λ^*F*^ | *Y, D*, **Z**].

Zhang and Tchetgen Tchetgen(2022) derived the specific form of the observed data efficient score for the considered model and introduced a *working model f*^*\**^(*U* | **Z**; *ξ*) instead of the unknown *f* (*U* | **Z**) to proceed calculation:

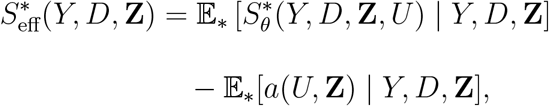

where *a*(*U*, **Z**) satisfies the integral equation:

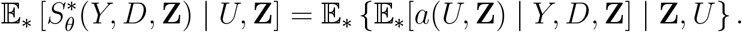

Note that the true data distribution *𝒫*_*F*_ can be factored as

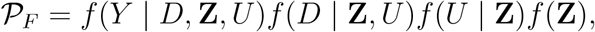

and the misspecified data distribution with working model is

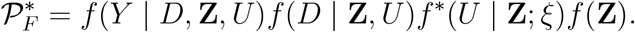

𝔼[*·*] denotes expectation taken with respect to *𝒫*_*F*_ and 𝔼_*\**_[*·*] taken with respect to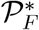. The efficient score 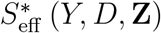 has an important property:

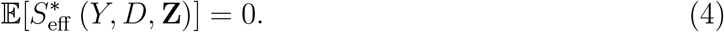

Equation (4) yields an estimator for *θ* that exhibits appealing robustness and efficiency properties. This is achieved by substituting the expectation with its empirical analogue and formulating the following estimating equation:

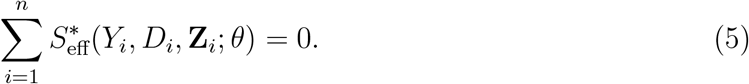

Under suitable identification and regularity conditions, the solution 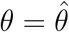 to the estimating equation (5) is consistent and asymptotically normal, with variance given by

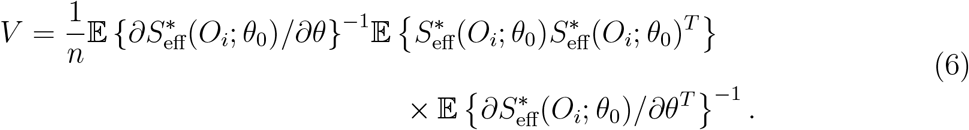

If the conditional distribution *f* (*U*|**Z**) is correctly specified, then 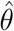 is locally efficient with asymptotic variance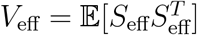.

### 2.4. Penalized Semiparametric Estimating Equation

Given estimating equation (5) and assumed identification condition, majority rule, we consider the penalized semiparametric estimating equation for simultaneous estimation and invalid instrumental variable selection. Specifically, the penalized semiparametric estimating functions are defined as

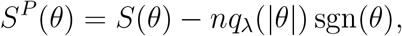

where 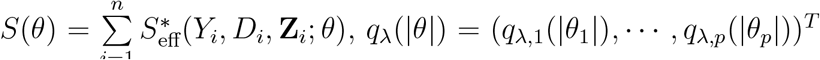 and the second term is the componentwise product of *q*_*λ*_ and sgn(*θ*). To select invalid instrumental variables, we design *q*_*λ*_(|*θ*|) as follows: (i) for *j* = 1, 2, *…, q* + 1, set *q*_*λ,j*_(|*θ*_*j*_|) = 0; (ii) for *j* = *q* + 2, *…, p*, set *q*_*λ,j*_(|*θ*_*j*_|) as SCAD penalty:

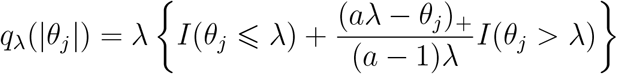

with *a >* 2.

Here we adopt the nonconvex SCAD penalty proposed by Fan and Li (2001), which results in an estimator with oracle property: that is, the estimator has the same limiting distribution as an estimator that konws the true model a priori.

Note that we only penalize the latter *q* component of *S*(*θ*) which corresponding to parameter *α*, therefore *γ* and *β* will not be shrunken. Intuitively, *q*_*λ*_(|*α*_*j*_|) is zero for a large value of |*α*_*j*_|, while it increases significantly for a small value of |*α*_*j*_|. Consequently, the jth component of the semiparametric estimating function *S*(*θ*), denoted as *S*_*j*_(*θ*), is not penalized when |*α*_*j*_| is large. Conversely, *S*_*j*_(*θ*) is heavily penalized if |*α*_*j*_| is close (but not equal) to zero, compelling the estimator of *α*_*j*_ to shrink to zero. When *α*_*j*_ is shrunk to zero, it implies that the jth instrumental variable is deemed valid and is consequently excluded from the outcome model.

## 3. Implementation and Results

### 3.1. Algorithm

To solve the penalized semiparametric estimating equation, we employ an iterative algorithm similar to that utilized in Johnson et al. (2008), Wang et al. (2012, 2013). This algorithm combines the minorization-maximization algorithm for the nonconvex penalty introduced by Hunter and Li (2005) with the Newton-Raphson algorithm:

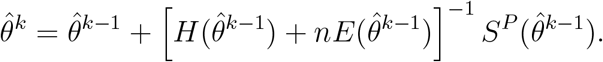

Where

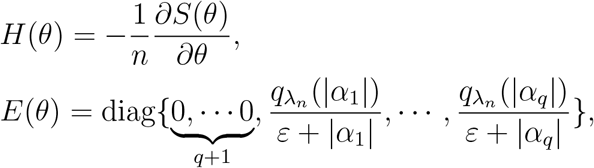

the constant *ϵ* represents a small perturbation, set to 10^*−*6^ in our simulation studies. We initialize the algorithm with the adaptive lasso estimator 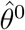. For a chosen tuning parameter, the algorithm iterates until the convergence criterion 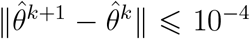 is satisfied. Typically, this criterion is met within 20 iterations in our simulation studies. Additionally, any coefficient that becomes sufficiently small is constrained to zero; specifically, if 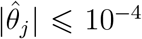 upon convergence, then the estimator for this coefficient is set to exactly zero.

We need to select (*a, λ*) for the SCAD penalty. Fan and Li (2001, 2002) demonstrated that the choice *a ≡* 3.7 performs well across various scenarios, and we adopt this recommendation for our numerical analyses. In practice, cross-validation is a widely used data-driven method for choosing *λ*. In the same vein, we employ *k*-fold cross-validation, minimizing the *L*_2_ norm of the estimating equation *S*(*θ*). This approach aligns with the fact that the parameter of interest is *θ*, which sets the expected value of the estimating equation to zero (see Equation (4)).

We derive the following sandwich formula from the algorithm to estimate the asymptotic covariance matrix of 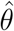:

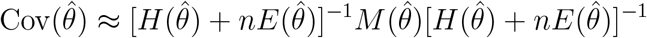

where 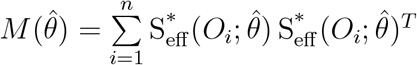

### 3.2. Asymptotic Theory for Penalized Semiparametric Estimator

Let 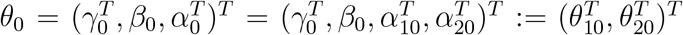 denote the true value of *θ*, where 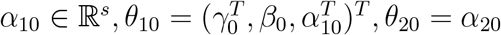 and 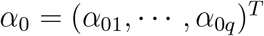. Without loss of generality, suppose that 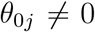 for *j* ⩽ *q* + 1 + *s* and 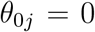 for *j > q* + 1 + *s*. For the asymptotic theory, we require the following regularity conditions.

a. 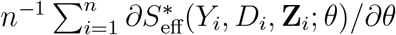 exists and is continuous in an open neighborhood of *β*_0_;
b. 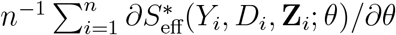 converges uniformly to its limit in a neighborhood of *θ*_0_;
c. 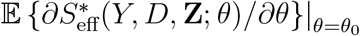 is invertible;
d. *λ*_*n*_ *→* 0 and 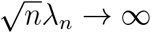

#### Remark 1

Conditions (a)-(c) are imposed by Zhang and Tchetgen Tchetgen (2022) to ensure the consistency and asymptotic normality of the estimator derived from the semiparametric estimating equation *S*(*θ*) = 0. Condition (d) represents a standard requirement concerning the rate of the tuning parameter to attain the oracle property (Fan and Li, 2001).

#### Theorem 1

Assuming conditions (a)-(d), the following results hold:

a. There exists a root-n-consistent approximate solution of 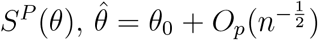, in the sense that 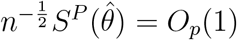.
b. (Oracle Property). For any root-n-consistent approximate solution 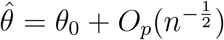, we have that 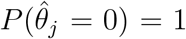 for *j > q* + 1 + *s*. Furthermore, if 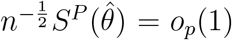, then 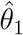 has the asymptotic normality

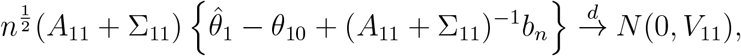

where *A*_11_, *V*_11_ are the first (*q* + 1 + *s*) *×* (*q* + 1 + *s*) submatrices of 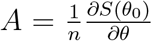 and *V* (as defined by Equation (6)), 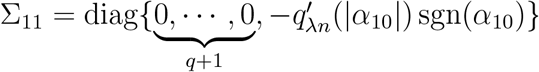, and

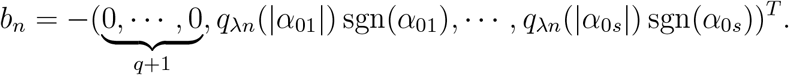
c. Let 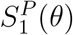 denote the first (*q* + 1 + *s*)-components of *S*^*P*^ (*θ*), then there exists 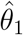 such that

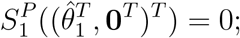

that is, the solution is exact.

## 4. Simulation Study

We consider a binary response *Y* and continuous exposure *D* in this section. Assume there are *n* = 1000 individuals and *q* = 10 candidate instruments. The observations (*Y*_*i*_, *D*_*i*_, **Z**_*i*_), *i* = 1, *…, n* are generated by

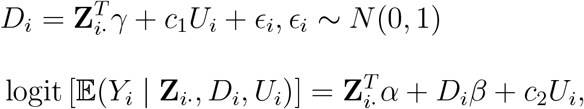

where **Z**_*i·*_ is drawn from a multivariate normal with zero mean and identity covariance matrix. We set *β* = 2, the fixed effect *γ* are drawn from *N* (0, 1). We vary (i) the direct effect parameter *α* = (1, 1, *…*, 0, 0) where we change *s* in ∥*α*∥_0_ = *s*, (ii) the distribution of unmeasured confounder *U* to test the robustness of our PSEE estimator.

Under each simulation scenario, we conduct 1000 replications. Our evaluation involves comparing the proposed PSEE method for estimating *β* with the original SEE method developed by Zhang and Tchetgen Tchetgen (2022). Additionally, we compute estimates from the “naive” Two-Stage Least Squares (TSLS) method under the assumption that all instruments are valid, and the “oracle” TSLS method, assuming perfect knowledge of which instruments are valid. Our focus is on the estimation accuracy and invalid instrument selection properties of these methods, assessed through bias and root mean square error (RMSE), along with the average number of correct (C) and incorrect (I) zero estimates, respectively. Additionally, we calculate the sample standard deviation of 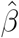 and the mean of the estimated standard deviation using the sandwich variance, denoted as SD_1_ and SD_2_. We also employ the sandwich variance formula to construct approximate 95% confidence intervals, relying on asymptotic normality theory, and report the corresponding empirical coverage probabilities.

The results of Table 1 summarize the performance of the naive TSLS, oracle TSLS, PSEE, and SEE for different number of invalid instruments *s*. The true distribution of unmeasured confounder *U* is Bernoulli(0.2) and *c*_1_ = *c*_2_ = 1. The PSEE (correct) and SEE (correct) denote estimators with correctly specified working model *U ∼* Bernoulli(0.2), the PSEE (incorrect) and SEE (incorrect) denote estimators with incorrectly specified working model *U ∼* Bernoulli(0.5). We observe that when majority rule holds, PSEE performs close to oracle TSLS in terms of instrument selection properties, and the estimated standard deviation closely approximates the empirical standard deviation, and the empirical coverage probability closely approaches 95%. These numerical results suggest the effective performance of the sandwich variance formula. Additionally, it is evident that PSEE outperforms naive TSLS and SEE in bias and RMSE, even when the majority rule is violated. We also observe that PSEE performs well with incorrectly specified working model, these indicate Equation (4).

**Table 1:** Estimation Results for *β* (*n* = 1000, *q* = 10, *U ∼* Bernoulli(0.2), *c*_1_ = *c*_2_ = 1): comparison of naive TSLS, oracle TSLS, PSEE (correct), SEE (correct), PSEE (incorrect) and SEE (incorrect). Here “correct” denotes estimator with correctly specified working model, i.e., *f*^*\**^(*U* |**Z**; *ξ*) = Bernoulli(0.2); “incorrect” denotes estimator with incorrectly specified working model, i.e., *f*^*\**^(*U* |**Z**; *ξ*) = Bernoulli(0.5). SD_1_ and SD_2_ denote the sample standard deviation and the mean of the estimated standard deviation using the sandwich variance. C and I denote the average number of correct and incorrect zero estimates, respectively. True *β* equals 2.0.

| | Bias | RMSE | $\text{SD}_1$ | $\text{SD}_2$ | Coverage | C | I |
| --- | --- | --- | --- | --- | --- | --- | --- |
| Majority rule holds |  |  |  |  |  |  |  |
| 1 invalid IV |  |  |  |  |  |  |  |
| PSEE(correct) | 0.053 | 0.180 | 0.162 | 0.172 | 93.4% | 8.644 | 0.026 |
| SEE(correct) | 0.065 | 0.231 | 0.216 | 0.222 | 95.1% |  |  |
| PSEE(incorrect) | 0.051 | 0.180 | 0.162 | 0.172 | 93.4% | 8.652 | 0.021 |
| SEE(incorrect) | 0.089 | 0.239 | 0.216 | 0.222 | 95.1% |  |  |
| Oracle | -0.008 | 0.161 |  |  |  | 9 | 0 |
| Naive | -0.814 | 0.817 |  |  |  |  |  |
| 3 invalid IVs |  |  |  |  |  |  |  |
| PSEE(correct) | 0.043 | 0.183 | 0.170 | 0.178 | 94.8% | 6.877 | 0.097 |
| SEE(correct) | 0.067 | 0.242 | 0.227 | 0.233 | 94.1% |  |  |
| PSEE(incorrect) | 0.042 | 0.183 | 0.171 | 0.178 | 94.4% | 6.868 | 0.078 |
| SEE(incorrect) | 0.090 | 0.250 | 0.227 | 0.233 | 93.6% |  |  |
| Oracle | -0.007 | 0.164 |  |  |  | 7 | 0 |
| Naive | -0.781 | 0.784 |  |  |  |  |  |
| Majority rule is violated |  |  |  |  |  |  |  |
| 5 invalid IVs |  |  |  |  |  |  |  |
| PSEE(correct) | 0.079 | 0.217 | 0.181 | 0.202 | 91.8% | 4.881 | 0.235 |
| SEE(correct) | 0.099 | 0.271 | 0.235 | 0.252 | 93.6% |  |  |
| PSEE(incorrect) | 0.078 | 0.216 | 0.180 | 0.202 | 91.8% | 4.883 | 0.268 |
| SEE(incorrect) | 0.122 | 0.281 | 0.235 | 0.253 | 93.3% |  |  |
| Oracle | 0.026 | 0.187 |  |  |  | 5 | 0 |
| Naive | -0.724 | 0.728 |  |  |  |  |  |
| 7 invalid IVs |  |  |  |  |  |  |  |
| PSEE(correct) | 0.072 | 0.221 | 0.181 | 0.208 | 91.7% | 2.951 | 0.756 |
| SEE(correct) | 0.081 | 0.263 | 0.230 | 0.251 | 93.6% |  |  |
| PSEE(incorrect) | 0.074 | 0.222 | 0.181 | 0.210 | 91.5% | 2.945 | 0.753 |
| SEE(incorrect) | 0.106 | 0.272 | 0.230 | 0.251 | 93.0% |  |  |
| Oracle | 0.066 | 0.208 |  |  |  | 3 | 0 |
| Naive | -0.901 | 0.903 |  |  |  |  |  |

Table 2 summarizes the performance when the unmeasured confounder is continuous, *U ∼ N* (0, 1), and *c*_1_ = *c*_2_ = 0.5. Our working model for *U* is a discrete uniform distribution on the interval [*−*0.5, 0.5] with mesh size *h*. For computationally efficiency, we take *h* = 0.5. We observe that when majority rule holds, PSEE performs close to oracle TSLS in terms of bias, RMSE and instrument selection properties, and coverage approaches 95%. When majority rule does not hold, PSEE also has good performance in bias and RMSE. Table 3 presents the results for an alternative continuous unmeasured confounder setting, *U ∼ t*(3), with *c*_1_ = *c*_2_ = 0.25. The working model is consistent with that of Table 2. It is concluded from Table 3 again that PSEE exhibits robust performance across various evaluation metrics in this scenario as well.

**Table 2:** Estimation Results for *β* (*n* = 1000, *q* = 10, *U∼ N* (0, 1), *c*_1_ = *c*_2_ = 0.25): comparison of naive TSLS, oracle TSLS, PSEE, SEE. Working model for *U* is a discrete uniform distribution on the interval [*−* 0.5, 0.5] with mesh size *h*, here we take *h* = 0.5. SD_1_ and SD_2_ denote the sample standard deviation and the mean of the estimated standard deviation using the sandwich variance. C and I denote the average number of correct and incorrect zero estimates, respectively. True *β* equals 2.0.

| | Bias | RMSE | $SD_1$ | $SD_2$ | Coverage | C | I |
| --- | --- | --- | --- | --- | --- | --- | --- |
| Majority rule holds |  |  |  |  |  |  |  |
| 1 invalid IV |  |  |  |  |  |  |  |
| PSEE | 0.015 | 0.168 | 0.159 | 0.168 | 93.9% | 8.694 | 0.005 |
| SEE | 0.105 | 0.245 | 0.215 | 0.221 | 94.4% |  |  |
| Oracle | 0.006 | 0.162 |  |  |  | 9 | 0 |
| Naive | -0.715 | 0.719 |  |  |  |  |  |
| 3 invalid IVs |  |  |  |  |  |  |  |
| PSEE | 0.033 | 0.173 | 0.167 | 0.170 | 95.0% | 6.942 | 0.018 |
| SEE | 0.128 | 0.259 | 0.221 | 0.225 | 94.1% |  |  |
| Oracle | 0.027 | 0.167 |  |  |  | 7 | 0 |
| Naive | -0.724 | 0.728 |  |  |  |  |  |
| Majority rule is violated |  |  |  |  |  |  |  |
| 5 invalid IVs |  |  |  |  |  |  |  |
| PSEE | 0.047 | 0.187 | 0.177 | 0.181 | 94.1% | 4.976 | 0.076 |
| SEE | 0.124 | 0.265 | 0.226 | 0.234 | 93.8% |  |  |
| Oracle | 0.048 | 0.183 |  |  |  | 5 | 0 |
| Naive | -0.641 | 0.646 |  |  |  |  |  |
| 7 invalid IVs |  |  |  |  |  |  |  |
| PSEE | 0.050 | 0.227 | 0.181 | 0.222 | 89.4% | 2.943 | 0.438 |
| SEE | 0.124 | 0.270 | 0.229 | 0.240 | 93.4% |  |  |
| Oracle | 0.059 | 0.204 |  |  |  | 3 | 0 |
| Naive | -0.940 | 0.942 |  |  |  |  |  |

**Table 3:** Estimation Results for *β* (*n* = 1000, *q* = 10, *U∼ t*(3), *c*_1_ = *c*_2_ = 0.25): comparison of naive TSLS, oracle TSLS, PSEE, SEE. Working model for *U* is a discrete uniform distribution on the interval [*−* 0.5, 0.5] with mesh size *h*, here we take *h* = 0.5. SD_1_ and SD_2_ denote the sample standard deviation and the mean of the estimated standard deviation using the sandwich variance. C and I denote the average number of correct and incorrect zero estimates, respectively. True *β* equals 2.0.

| | Bias | RMSE | $SD_1$ | $SD_2$ | Coverage | C | I |
| --- | --- | --- | --- | --- | --- | --- | --- |
| Majority rule holds |  |  |  |  |  |  |  |
| 1 invalid IV |  |  |  |  |  |  |  |
| PSEE | 0.004 | 0.160 | 0.155 | 0.160 | 94.1% | 8.633 | 0.014 |
| SEE | 0.163 | 0.268 | 0.208 | 0.213 | 90.4% |  |  |
| Oracle | -0.010 | 0.155 |  |  |  | 9 | 0 |
| Naive | -0.780 | 0.783 |  |  |  |  |  |
| 3 invalid IVs |  |  |  |  |  |  |  |
| PSEE | 0.023 | 0.186 | 0.168 | 0.185 | 94.0% | 6.886 | 0.040 |
| SEE | 0.182 | 0.297 | 0.224 | 0.235 | 89.6% |  |  |
| Oracle | 0.014 | 0.179 |  |  |  | 7 | 0 |
| Naive | -0.782 | 0.785 |  |  |  |  |  |
| Majority rule is violated |  |  |  |  |  |  |  |
| 5 invalid IVs |  |  |  |  |  |  |  |
| PSEE | 0.049 | 0.191 | 0.175 | 0.185 | 94.5% | 4.927 | 0.119 |
| SEE | 0.187 | 0.299 | 0.226 | 0.233 | 91.5% |  |  |
| Oracle | 0.049 | 0.191 |  |  |  | 5 | 0 |
| Naive | -0.633 | 0.638 |  |  |  |  |  |
| 7 invalid IVs |  |  |  |  |  |  |  |
| PSEE | 0.061 | 0.220 | 0.179 | 0.212 | 91.5% | 2.905 | 0.599 |
| SEE | 0.195 | 0.310 | 0.231 | 0.241 | 89.5% |  |  |
| Oracle | 0.065 | 0.206 |  |  |  | 3 | 0 |
| Naive | -0.917 | 0.919 |  |  |  |  |  |

## 5. Real Data Analysis

We demonstrate the potential advantages of our method in Mendelian randomization (MR) by analyzing the effect of BMI on suffering stroke. For this analysis, we leverage data from the Atherosclerosis Risk in Communities Study (ARIC), which is a prospective longitudinal epidemiological study conducted in four U.S. communities in North Carolina, Massachusetts, Maryland, and Minnesota.

Similar to another analysis with the ARIC data, we include individuals of white origin and extract European ancestry individuals and impute the data set on Michigan Imputation Center with EUR population from 1000 Genomes Phase 3 v5 reference panel (Shi et al., 2023). We remove 0.82% missing data in the following analysis. Finally, 8739 individuals are selected.

We consider potential candidate instruments for our MR analysis using the following SNPs in the ARIC data that have been previously associated with BMI: rs725959, rs1147199, rs3817334, and rs6477694. While we have no specific reason to believe any of these SNPs are invalid IVs, uncertainty arises due to incomplete knowledge about their biological functions. Additionally, the inability to control all confounders precisely is a common scenario in MR studies.

Under the assumption that all instruments are valid, the TSLS method estimates a causal effect of 0.0805 (OR = 1.0838). In contrast, PSEE (with working model *U ∼* Bernoulli(0.5)) estimates a causal effect of 0.1107 (SE: 0.0265, OR = 1.1171) with a 95% confidence interval [0.0588, 0.1626], excluding 0. The difference between TSLS and PSEE may stem from the underlying distribution of unmeasured confounder, as demonstrated in our simulations. Importantly, PSEE does not identify any SNPs as invalid IVs.

To further validate our method, we introduce another instrument, rs42039, associated with both BMI and stroke. Under the assumption that all four instruments are valid, TSLS estimates an effect of -0.0331 (OR = 0.9674). Conversely, PSEE (with working model *U ∼* Bernoulli(0.5)) estimates a causal effect of 0.1108 (SE: 0.0265, OR = 1.1171), similar to the estimates when using four instruments. PSEE also excludes rs42039, suspected to be invalid.

In the real data analysis, PSEE provides similar estimates and consistently excludes the suspected invalid instrument (rs42039) when additional instrument is introduced, this indicates its robustness to possibly invalid instruments compared to TSLS. Furthermore, Harshfield et al. (2021) found that heightened obesity will increase the risk of ischemic, large artery, and small vessel stroke, which supports our PSEE results. Therefore, it is recommended to focus on interventions that reduce obesity to mitigate the risk of stroke.

## 6. Discussions

In this paper, we consider a flexible semiparametric instrumental variable model accommodating for continuous/binary exposure/outcome. We assume identifiability of this model under majority rule and propose a penalized semiparametric approach to estimate causal effect. The asymptotic and oracle properties are outlined in Theorem 1 and further illustrated in comprehensive simulation studies. Specifically, our proposed method, PSEE, demonstrates performance close to that of the oracle TSLS regarding bias, RMSE, and instrument selection properties for both binary and continuous unmeasured confounder scenarios when majority rule holds. Even when majority rule does not hold, PSEE performs well in terms of bias and RMSE. Additionally, the robustness of our estimator is evident in both simulation experiments and real data analysis. We emphasize that PSEE does not impose specific requirements on whether the outcome and exposure are continuous or binary; it only requires the ability to specify the conditional probability distributions of the outcome and exposure. However, in our simulations, we specifically consider scenarios with binary outcome and continuous exposure. Nevertheless, it is worth noting that PSEE exhibits a lower coverage for increased values of sensitivity parameters *c*_1_ and *c*_2_, as illustrated in the Appendix C.

Further work could involve extending the considered model to the presence of additional complexities, such as nonlinear (Staley and Burgess, 2017) or time-varying exposure (Labrecque and Swanson, 2019), and vector-valued confounder. One could also consider different penalty functions, such as MCP (Zhang, 2010) and ALASSO (Zou, 2006) penalty, to compare their estimation accuracy and instrument selection properties. Moreover, the computational challenges posed by a considerable number of instrumental variables need to be tackled in future research. In our simulation experiments, involving a sample size of 1000 and 10 instrumental variables, the task of conducting 1000 replications on an 80-node cluster required an average of 30 hours. This highlights the necessity of developing efficient algorithms or optimization techniques, which would significantly enhance the scalability and practical applicability of our proposed method.

## Data Availability

All data produced are available online at https://www.ncbi.nlm.nih.gov/projects/gap/cgi-bin/study.cgi?study_id=phs000090.v1.p1

https://www.ebi.ac.uk/gwas/

## Acknowledgements

We express our gratitude to ARIC for generously providing the data. This research was supported partially by the National Key R&D Program of China [2023YFF1205101].

## Appendix

### Appendix A: Proof of Theorem 1

Proof: To prove part *a*, we consider 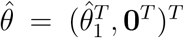, where 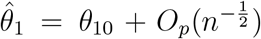. For *j* = 1, 2, *…, p*, we have

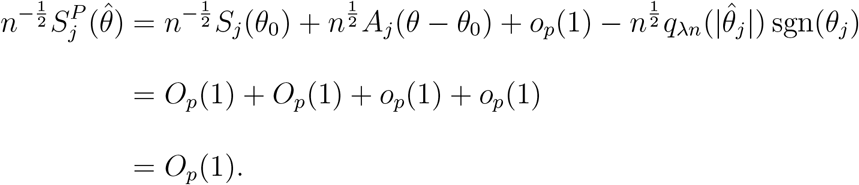

where *A*_*j*_ is the *j*^th^ row of *A*.

To prove part *b*, we consider the sets in the probability 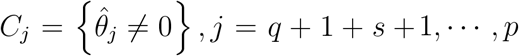. We show that for any *ε >* 0, when *n* is sufficiently large, *P* (*C*_*j*_) *< ε*. Because 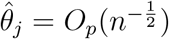, there exists some *M* such that when *n* is large enough,

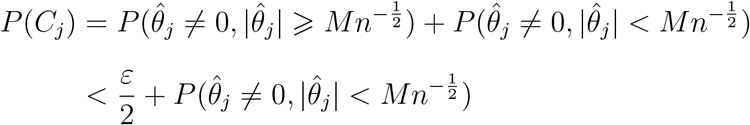

Using the jth component of the penalized estimating function and the definition of the approximate solution, we obtain that on the set of 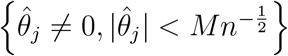,

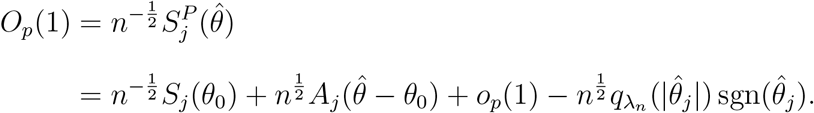

The first three terms on the right side are of order *O*_*p*_(1). As a result, there exists some *M*^*′*^ such that for large *n*,

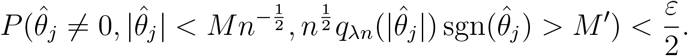

Because 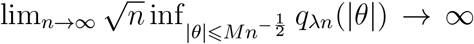 by condition 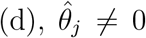 and 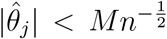 imply that 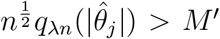 for large *n*. Thus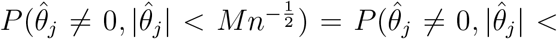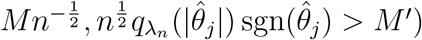. Therefore,

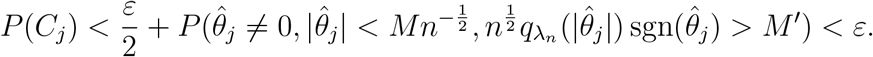

We next show the asymptotic normality of 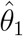 when the order of 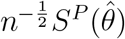 is *o*_*p*_(1). We have

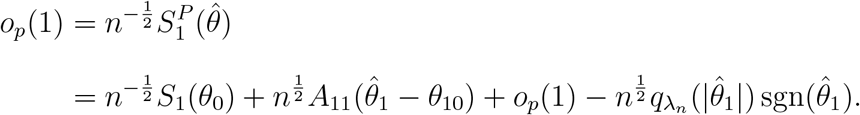

Employing Taylor expansions of 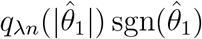 at *θ*_10_ yields

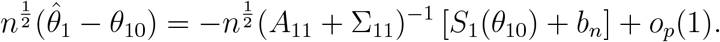

Where

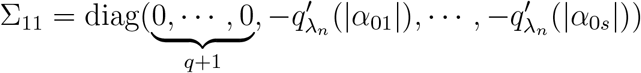

And

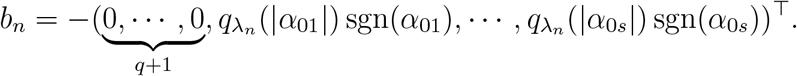

we then obtain that

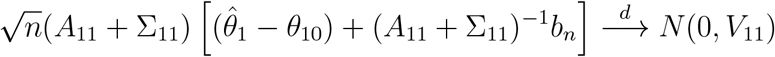

where *V*_11_ is the (*q* + 1 + *s*) *×* (*q* + 1 + *s*) submatrix in the upper-left corner of *V*. This completes the proof.

To establish part c, we examine *θ*_1_ ∈ ℝ^*q*+1+*s*^ situated on the boundary of a ball centered around *θ*_10_, defined as *θ*_1_ = *θ*_10_ + *n*^*−*1*/*2^*u* with |*u*| = *r* for a constant *r*. Leveraging the penalized estimating function 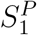, we derive the following expression:

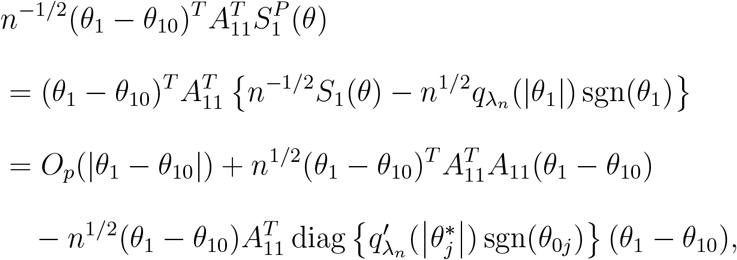

where 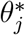 lies between *θ*_*j*_ and *θ*_0*j*_ for *j* = 1, *…, s*. As *A*_11_ is nonsingular, the second term on the right side exceeds *a*_0_*r*^2^*n*^*−*1*/*2^, where *a*_0_ denotes the smallest eigenvalue of 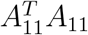. The first term is of order *rO*_*p*_(*n*^*−*1*/*2^). Due to the convergence of 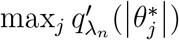 to 0, the third term is dominated by the second term. Therefore, by selecting *r* adequately large such that, for large *n*, the probability that the absolute value of the first term surpasses the second term is less than *ϵ*, we obtain

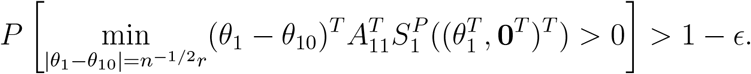

Applying the Brouwer fixed-point theorem to the continuous function 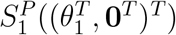, we see that 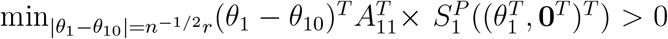 implies that 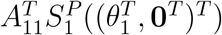 has a solution within this ball. In other words, 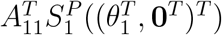 has a solution within this ball, or equivalently, 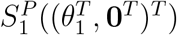 has a solution within this ball. Thus, we can select an exact solution 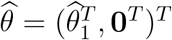 to 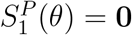 with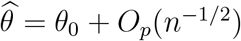.

### Appendix B: Boxplots and histograms for simulation studies

*U ∼ Bernoulli(0.2)*.

**Figure A1:**
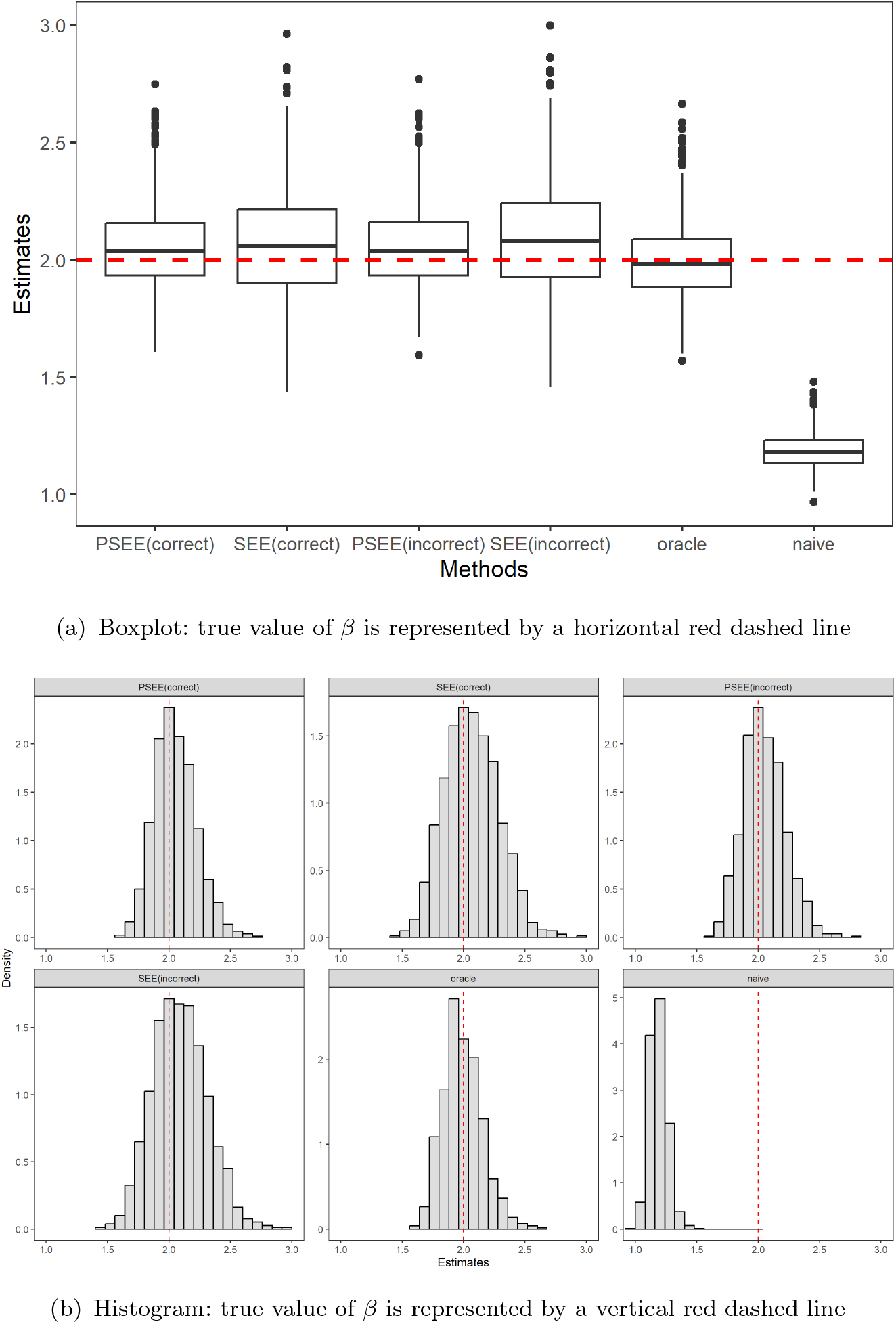
Boxplot and histogram of estimates of the causal effect *β, n = 1000, q =* 10, *U ∼* Bernoulli (0.2), *c*_1_ *= c*_2_ *=* 1, 1 invalid IV, 1000 replications.

**Figure A2:**
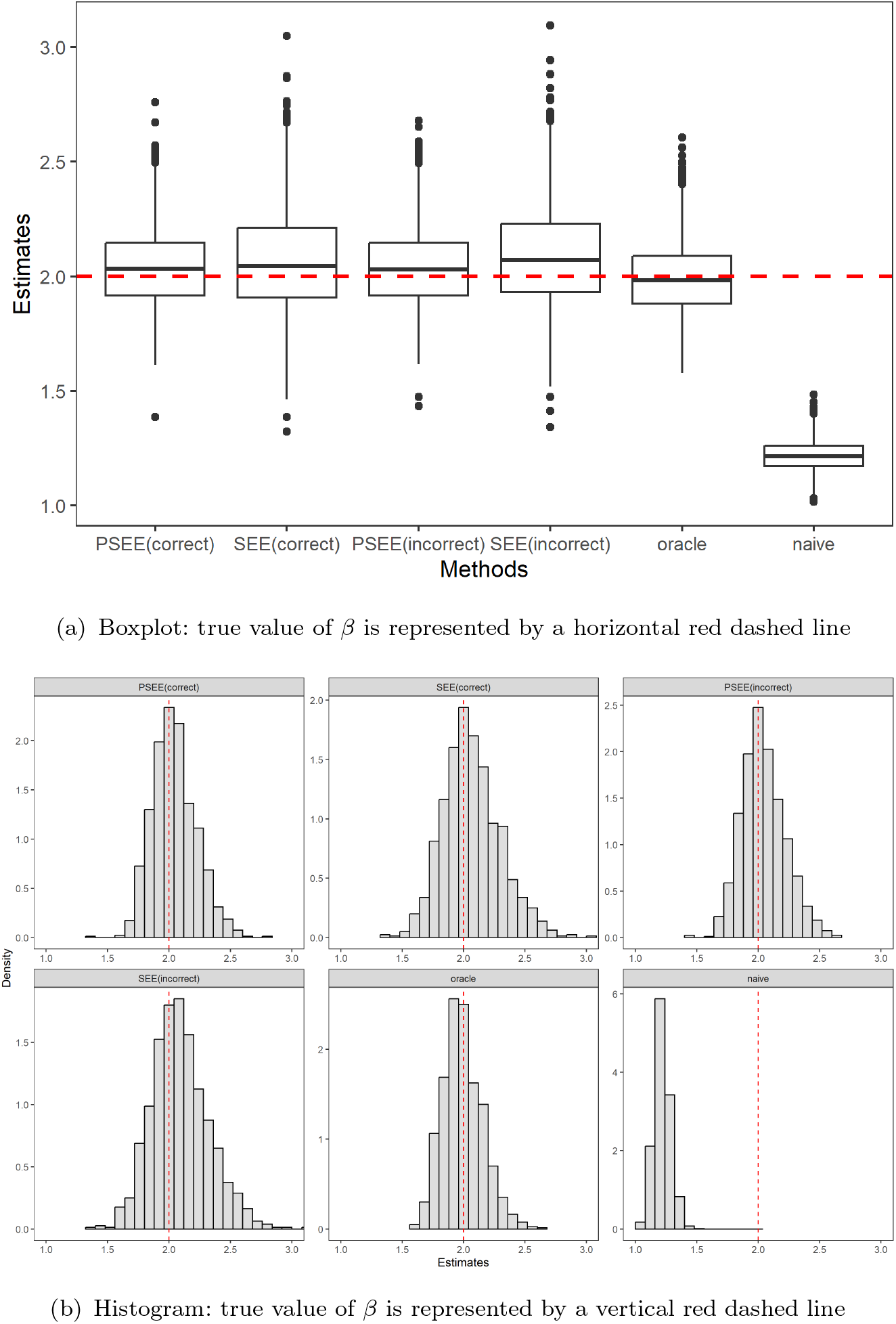
Boxplot and histogram of estimates of the causal effect *β, n =* 1000, *q =* 10, *U ∼* Bernoulli (0.2), *c*_1_ *= c*_2_ *=* 1, 3 invalid IVs, 1000 replications.

**Figure A3:**
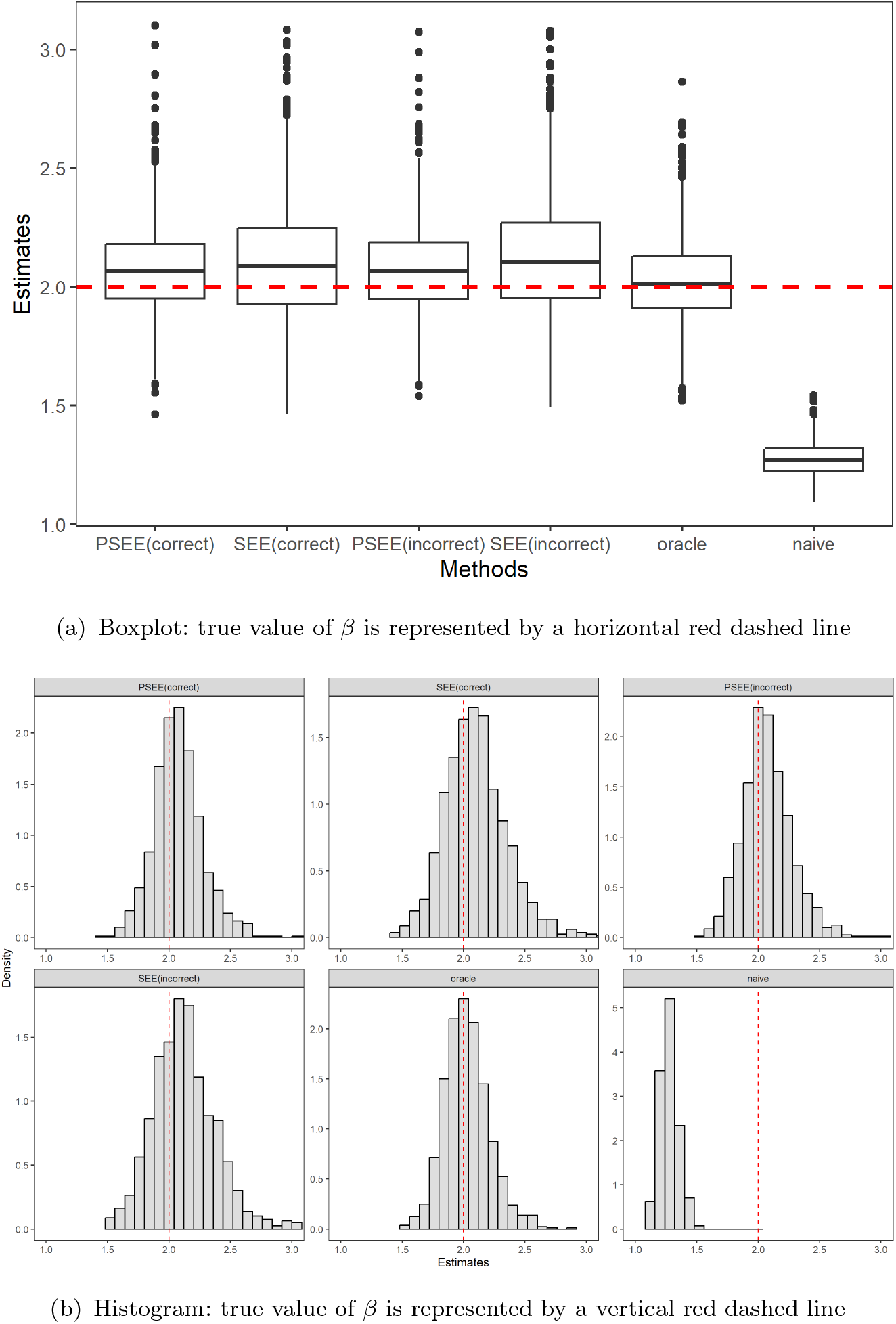
Boxplot and histogram of estimates of the causal effect *β, n* = 1000, q = 10, *U ∼* Bernoulli(0.2), *c*_1_ = *c*_2_ = 1, 5 invalid IVs, 1000 replications.

**Figure A4:**
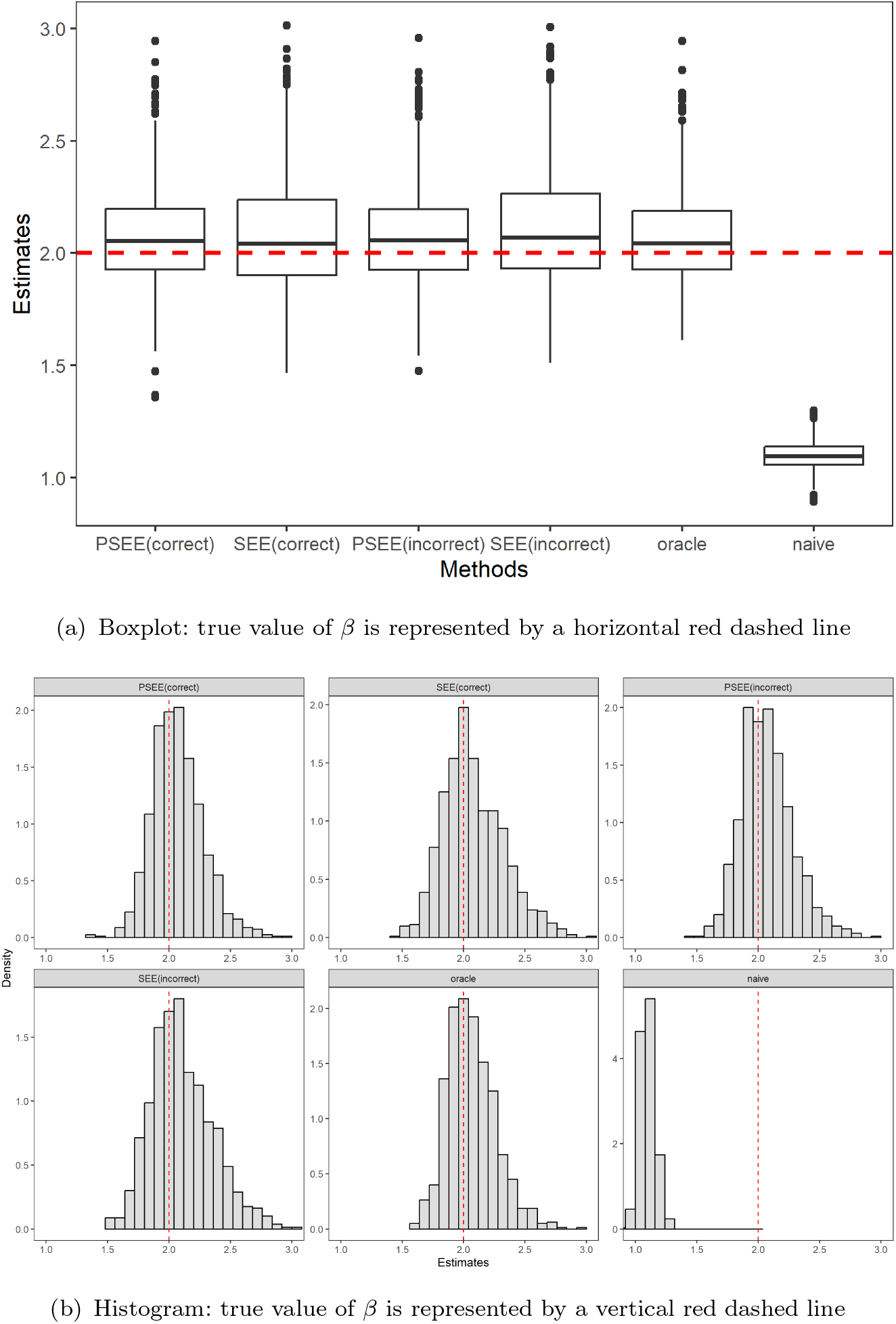
Boxplot and histogram of estimates of the causal effect *β, n* = 1000, *q* = 10, *U* ∼ Bernoulli(0.2), *c*_1_ = *c*_2_ = 1, 7 invalid IVs, 1000 replications.

*U ∼ N* (0, 1).

**Figure A5:**
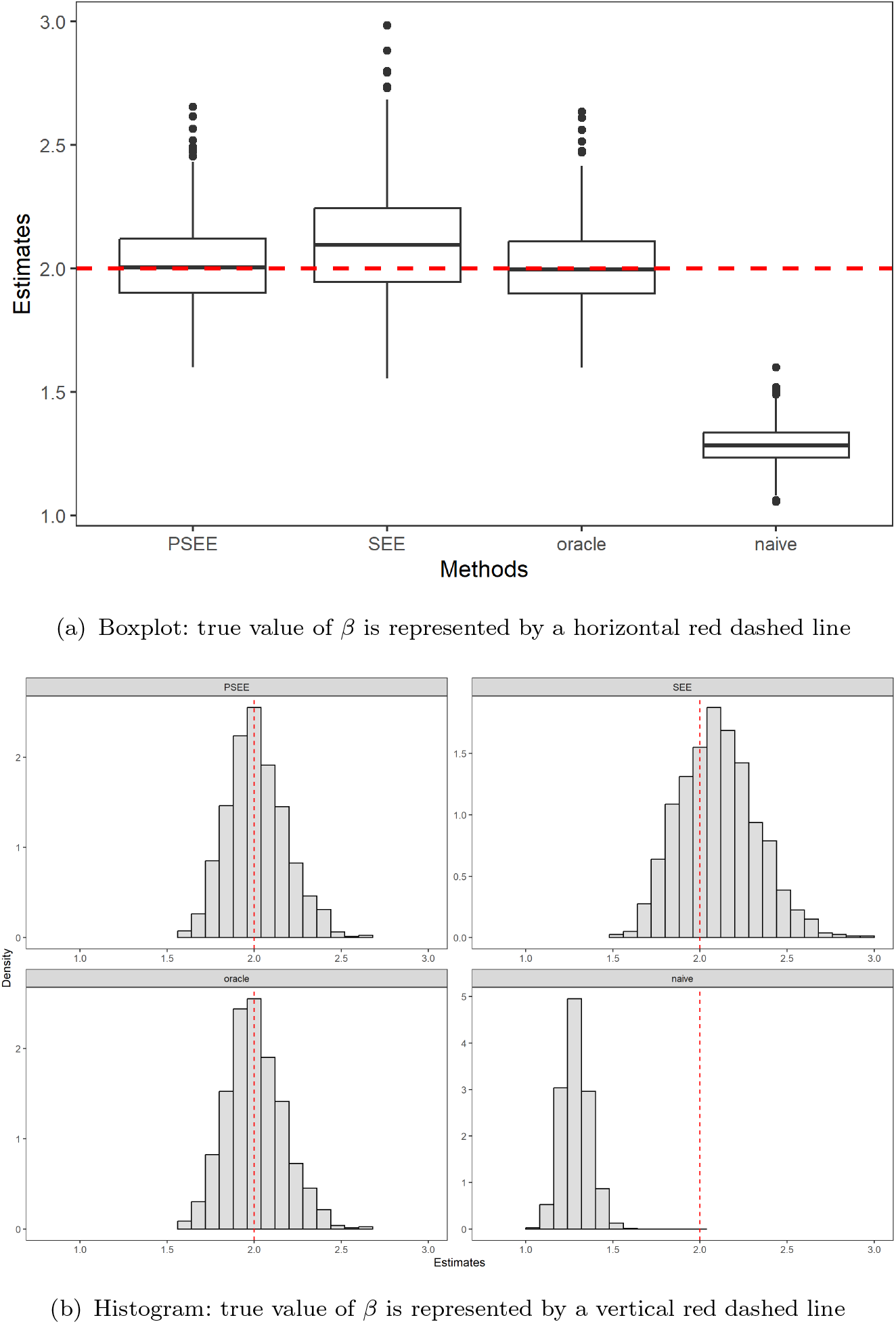
Boxplot and histogram of estimates of the causal effect *β, n* = 1000, *q* = 10, *U* ∼ *N* (0, 1), *c*_1_ = *c*_2_ = 0.25, 1 invalid IV, 1000 replications.

**Figure A6:**
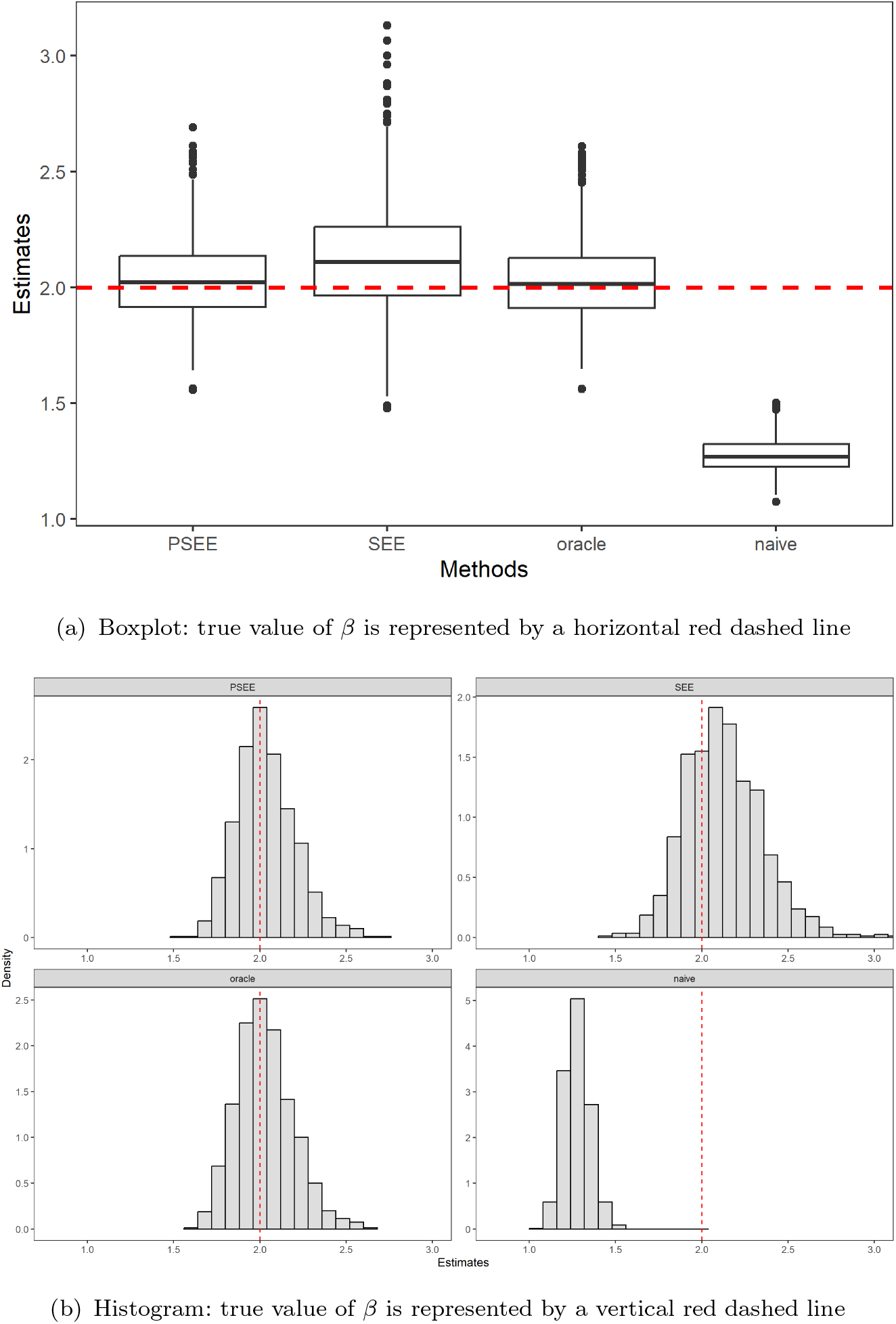
Boxplot and histogram of estimates of the causal effect *β, n* = 1000, *q* = 10, *U* ∼ *N* (0, 1), *c*_1_ = *c*_2_ = 0.25, 3 invalid IVs, 1000 replications.

**Figure A7:**
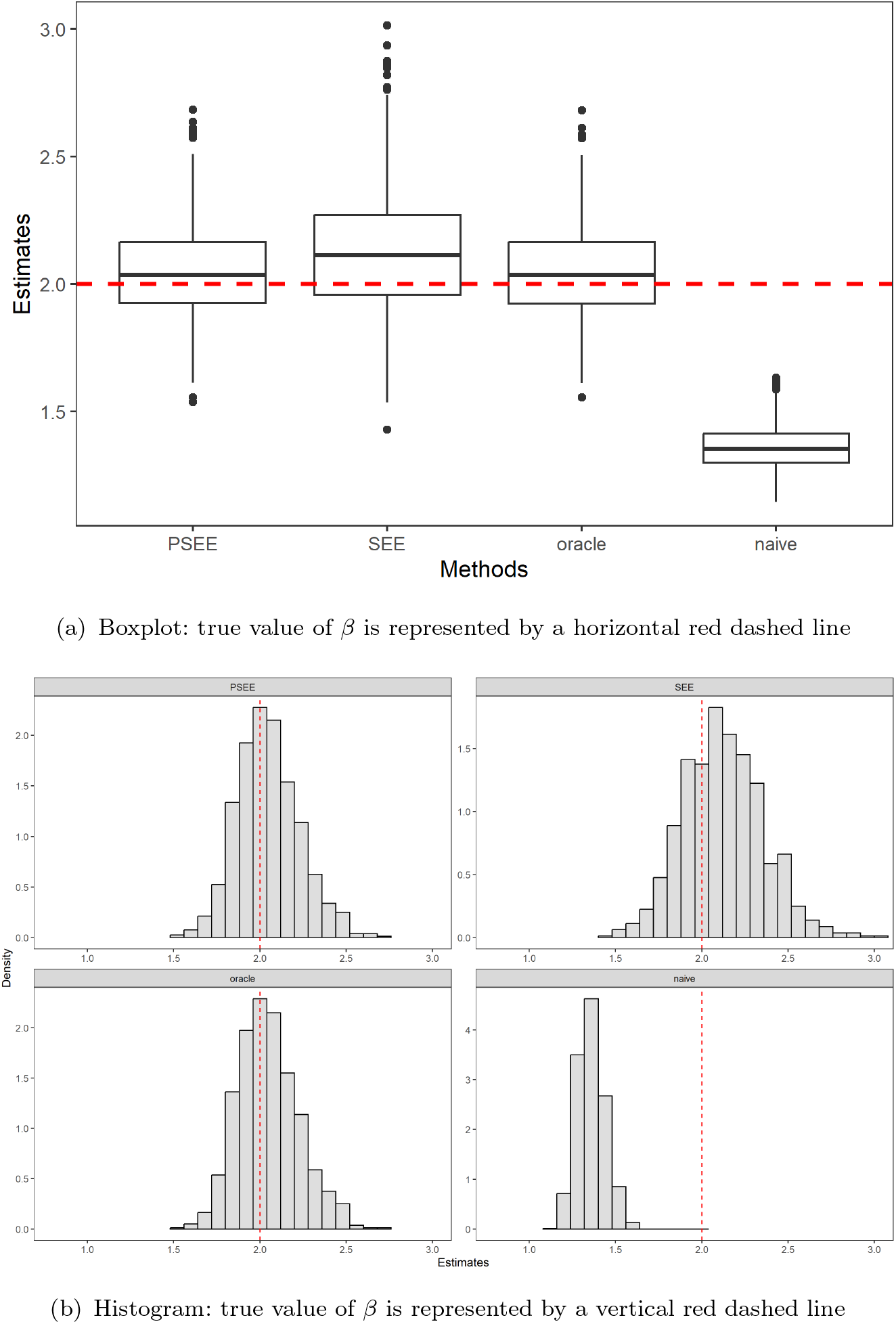
Boxplot and histogram of estimates of the causal effect *β, n* = 1000, *q* = 10, *U* ∼ *N* (0, 1), *c*_1_ = *c*_2_ = 0.25, 5 invalid IVs, 1000 replications.

**Figure A8:**
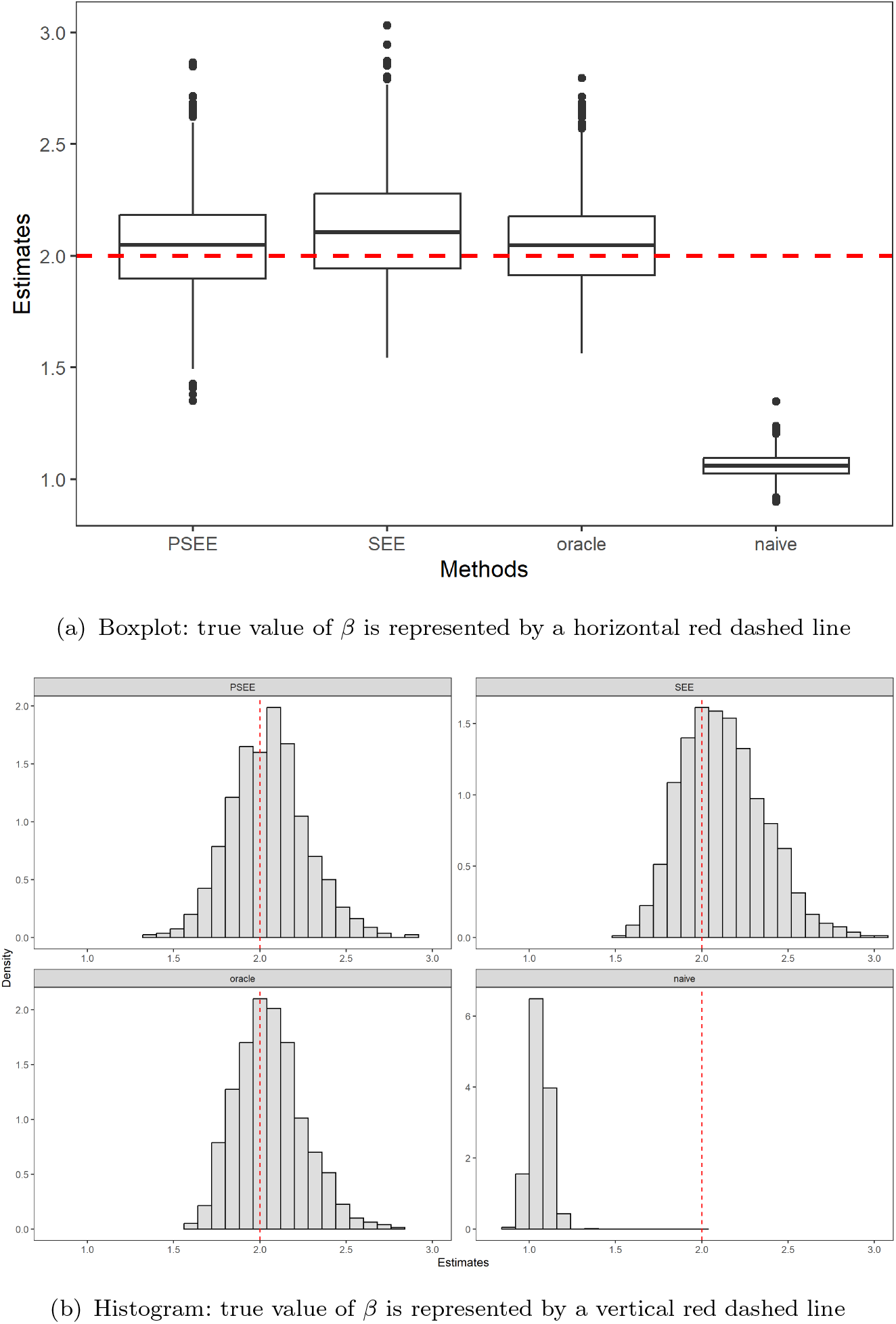
Boxplot and histogram of estimates of the causal effect *β, n* = 1000, *q* = 10, *U* ∼ *N*(0, 1), *c*_1_ = *c*_2_ = 0.25, 7 invalid IVs, 1000 replications.

*U ∼ t*(3).

**Figure A9:**
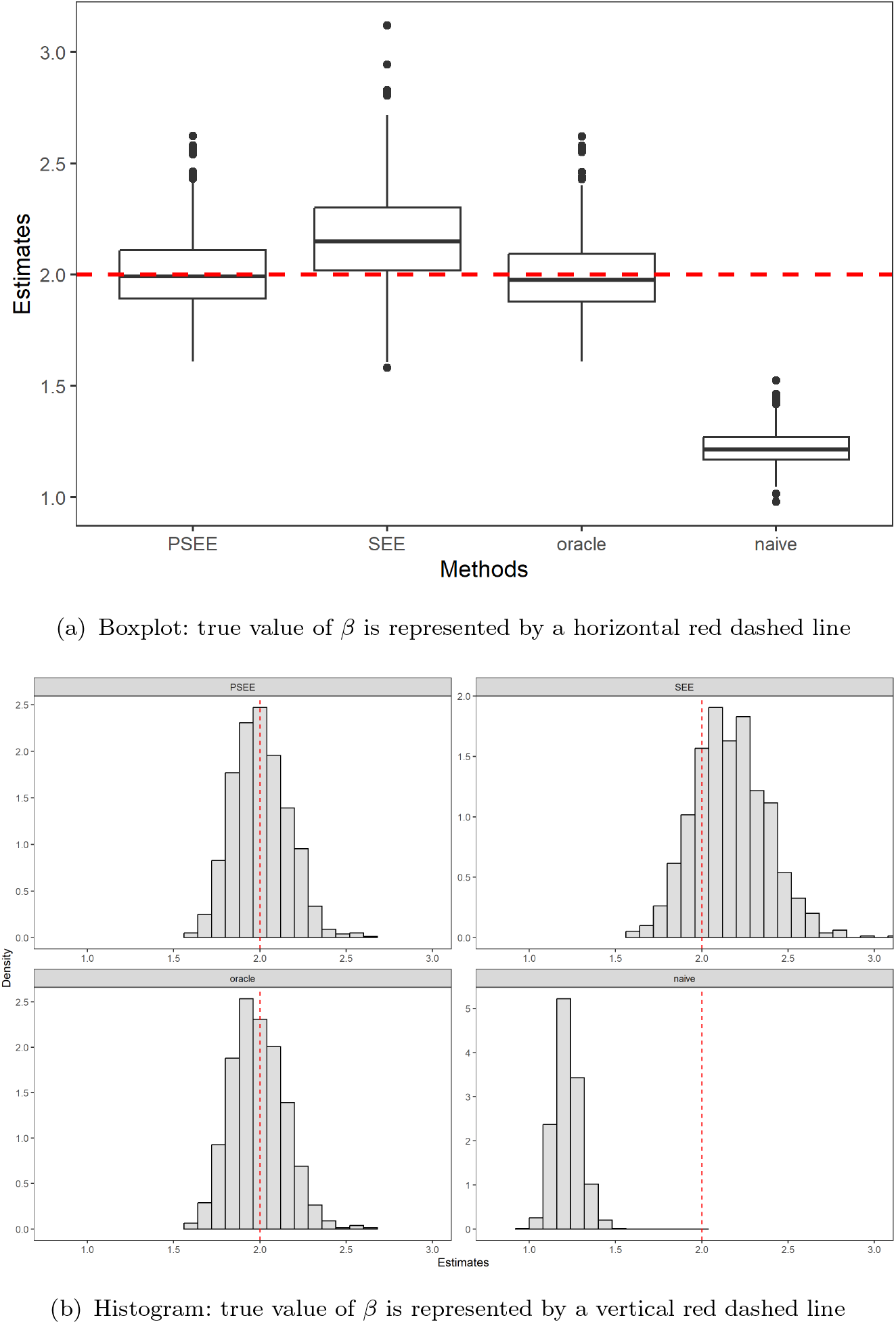
Boxplot and histogram of estimates of the causal effect *β, n* = 1000, *q* = 10, *U* ∼ *t* (3), *c*_1_ = *c*_2_ = 0.25, 1 invalid IV, 1000 replications.

**Figure A10:**
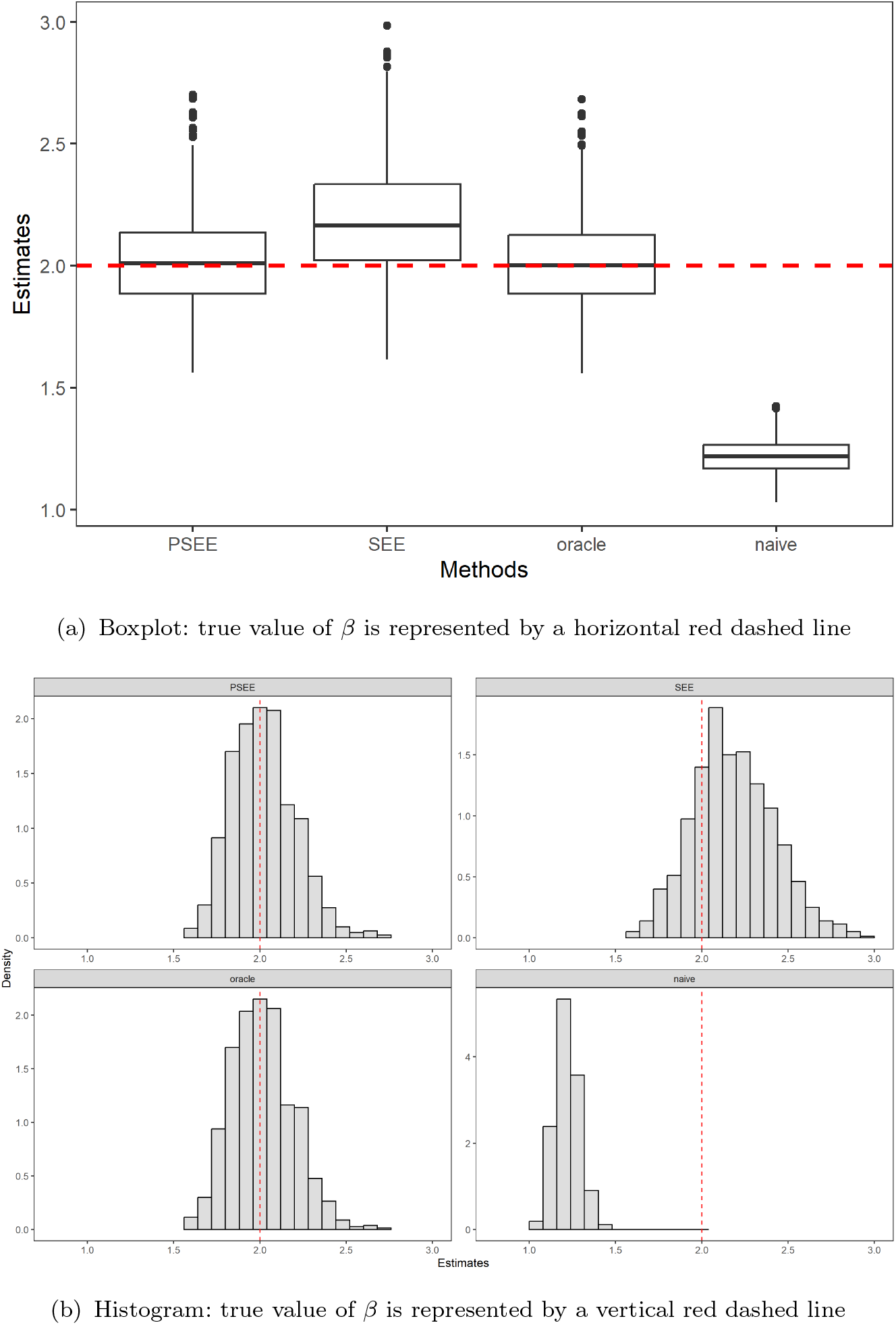
Boxplot and histogram of estimates of the causal effect *β, n* = 1000, *q* = 10, *U* ∼ *t*(3), *c*_1_ = *c*_2_ = 0.25, 3 invalid IVs, 1000 replications.

**Figure A11:**
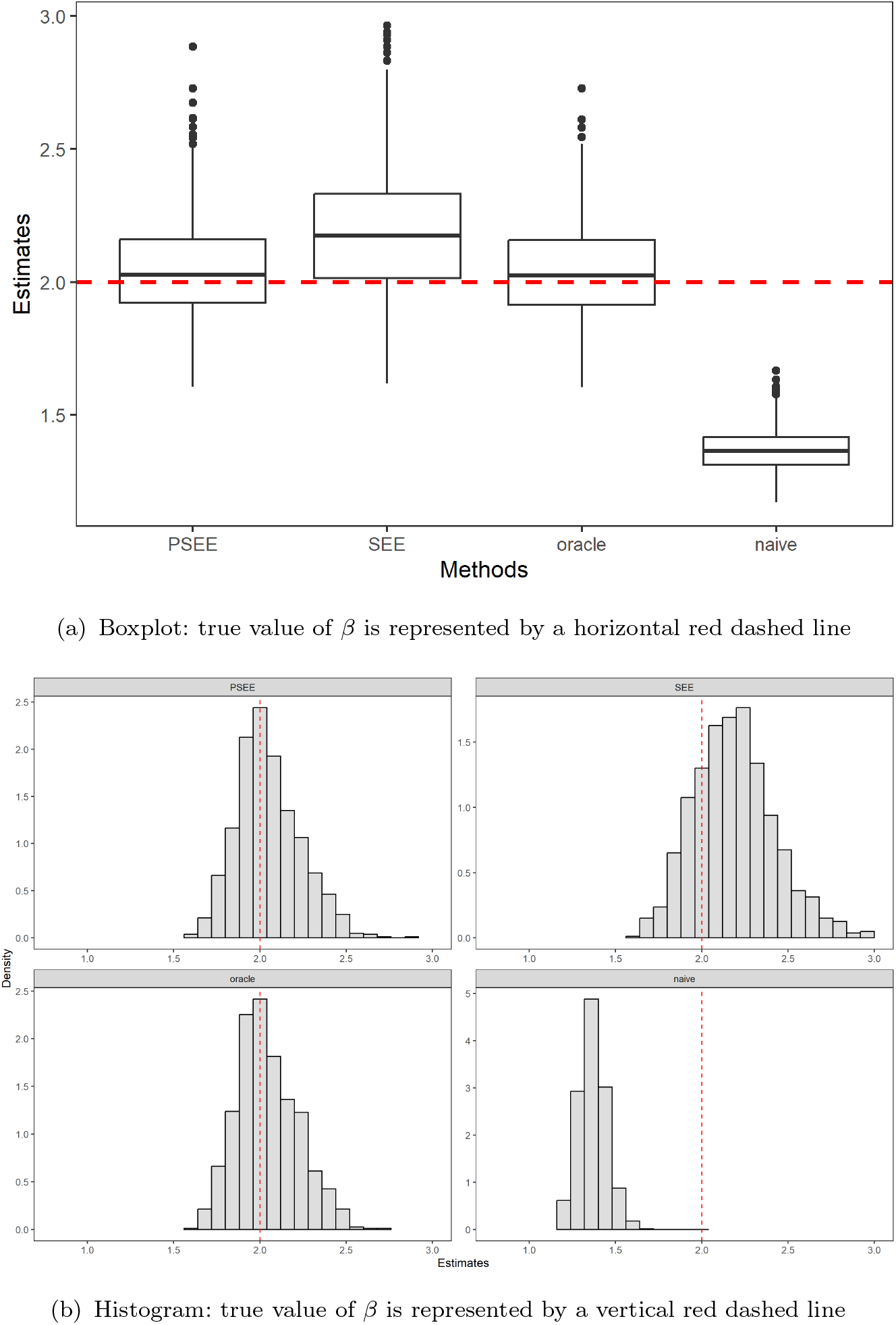
Boxplot and histogram of estimates of the causal effect *β, n* = 1000, *q* = 10, *U* ∼ *t*(3), *c*_1_ = *c*_2_ = 0.25, 5 invalid IVs, 1000 replications.

**Figure A12:**
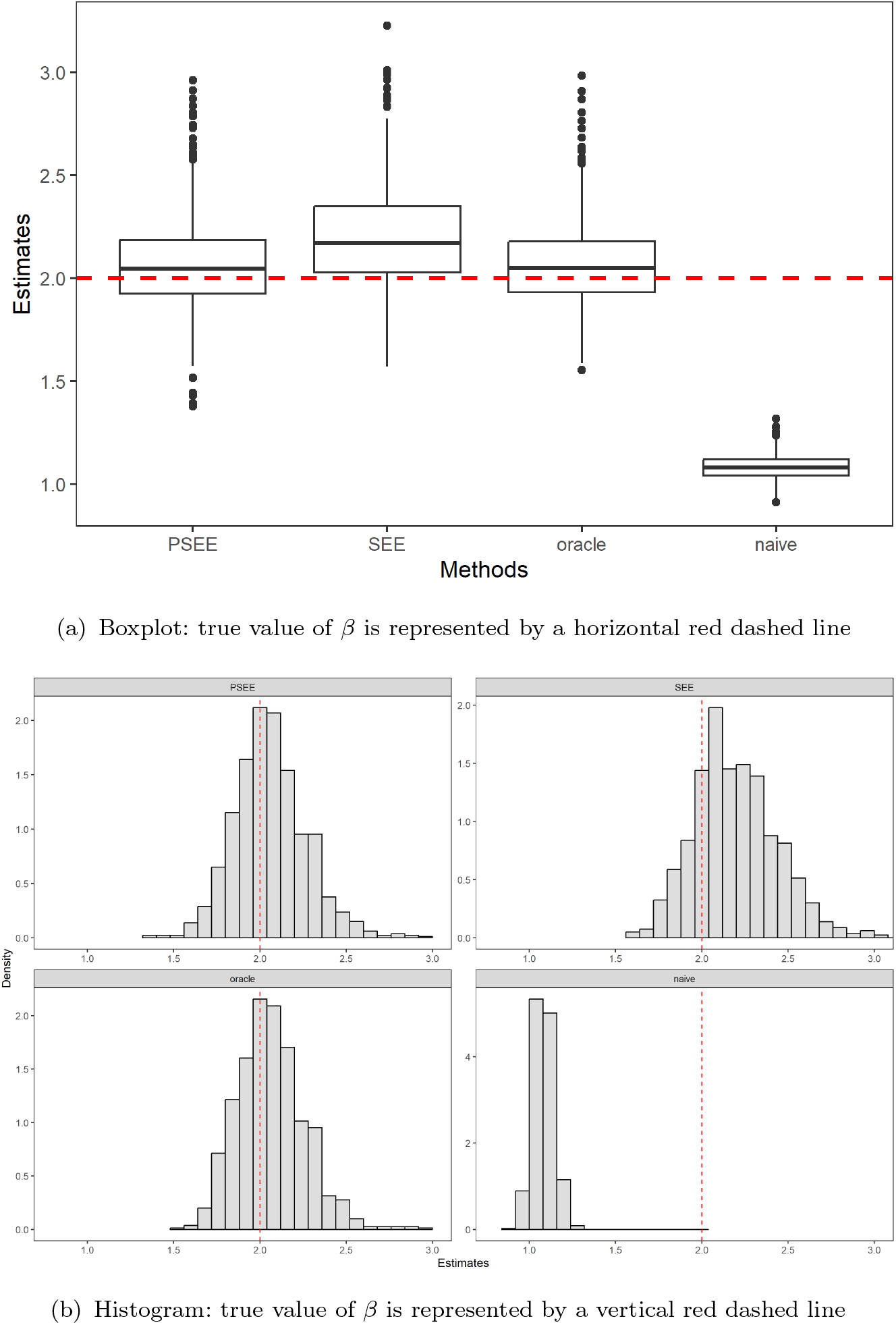
Boxplot and histogram of estimates of the causal effect *β, n* = 1000, *q* = 10, *U* ∼ *t*(3), *c*_1_ = *c*_2_ = 0.25, 7 invalid IVs, 1000 replications.

### Appendix C: Simulation results for increased values of sensitivity parameters

*U ∼ N* (0, 1) *and c*_1_ = *c*_2_ = 0.5.

**Table A1:** Estimation Results for *β* (*n* = 1000, *q* = 10, *U ∼ N* (0, 1), *c*_1_ = *c*_2_ = 0.5): comparison of naive TSLS, oracle TSLS, PSEE, SEE. Working model for *U* is a discrete uniform distribution on the interval [*−* 0.5, 0.5] with mesh size *h*, here we take *h* = 0.5. SD_1_ and SD_2_ denote the sample standard deviation and the mean of the estimated standard deviation using the sandwich variance. C and I denote the average number of correct and incorrect zero estimates, respectively. True *β* equals 2.0.

| | Bias | RMSE | $SD_1$ | $SD_2$ | Coverage | C | I |
| --- | --- | --- | --- | --- | --- | --- | --- |
| Majority rule holds |  |  |  |  |  |  |  |
| 1 invalid IV |  |  |  |  |  |  |  |
| PSEE | -0.005 | 0.165 | 0.158 | 0.165 | 94.9% | 8.683 | 0.024 |
| SEE | 0.201 | 0.300 | 0.217 | 0.223 | 87.9% |  |  |
| Oracle | -0.023 | 0.161 |  |  |  | 9 | 0 |
| Naive | -0.834 | 0.837 |  |  |  |  |  |
| 3 invalid IVs |  |  |  |  |  |  |  |
| PSEE | 0.005 | 0.180 | 0.163 | 0.180 | 92.4% | 6.878 | 0.079 |
| SEE | 0.194 | 0.299 | 0.219 | 0.227 | 88.1% |  |  |
| Oracle | -0.006 | 0.169 |  |  |  | 7 | 0 |
| Naive | -0.830 | 0.833 |  |  |  |  |  |
| Majority rule is violated |  |  |  |  |  |  |  |
| 5 invalid IVs |  |  |  |  |  |  |  |
| PSEE | 0.045 | 0.210 | 0.180 | 0.206 | 92.2% | 4.858 | 0.265 |
| SEE | 0.208 | 0.322 | 0.234 | 0.246 | 89.1% |  |  |
| Oracle | 0.029 | 0.194 |  |  |  | 5 | 0 |
| Naive | -0.667 | 0.672 |  |  |  |  |  |
| 7 invalid IVs |  |  |  |  |  |  |  |
| PSEE | 0.065 | 0.217 | 0.176 | 0.207 | 91.4% | 2.908 | 0.973 |
| SEE | 0.207 | 0.320 | 0.227 | 0.244 | 86.9% |  |  |
| Oracle | 0.057 | 0.202 |  |  |  | 3 | 0 |
| Naive | -0.983 | 0.984 |  |  |  |  |  |

**Figure A13:**
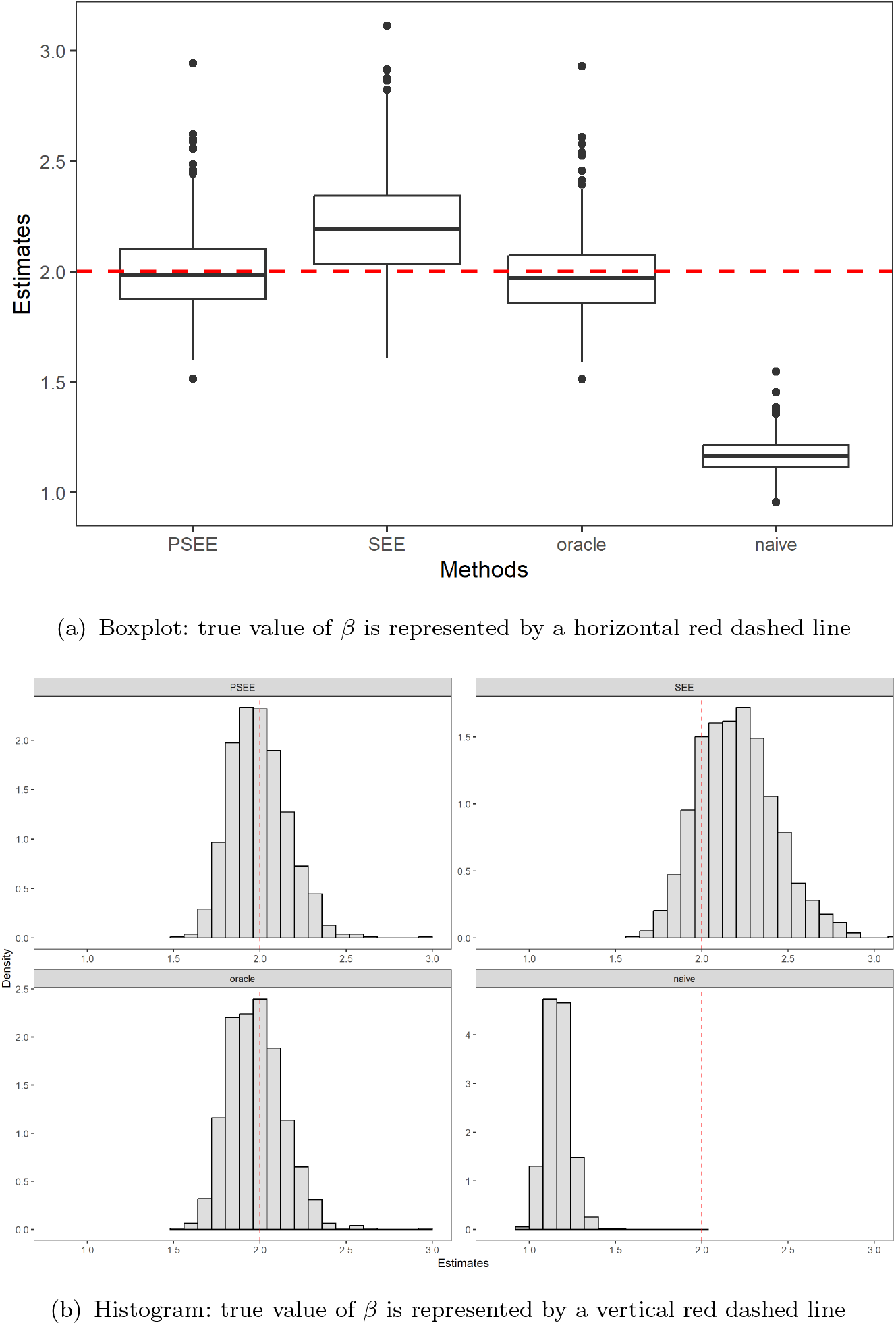
Boxplot and histogram of estimates of the causal effect *β, n* = 1000, *q* = 10, *U* ∼ *N*(0, 1), *c*_1_ = *c*_2_ = 0.5, 1 invalid IV, 1000 replications.

**Figure A14:**
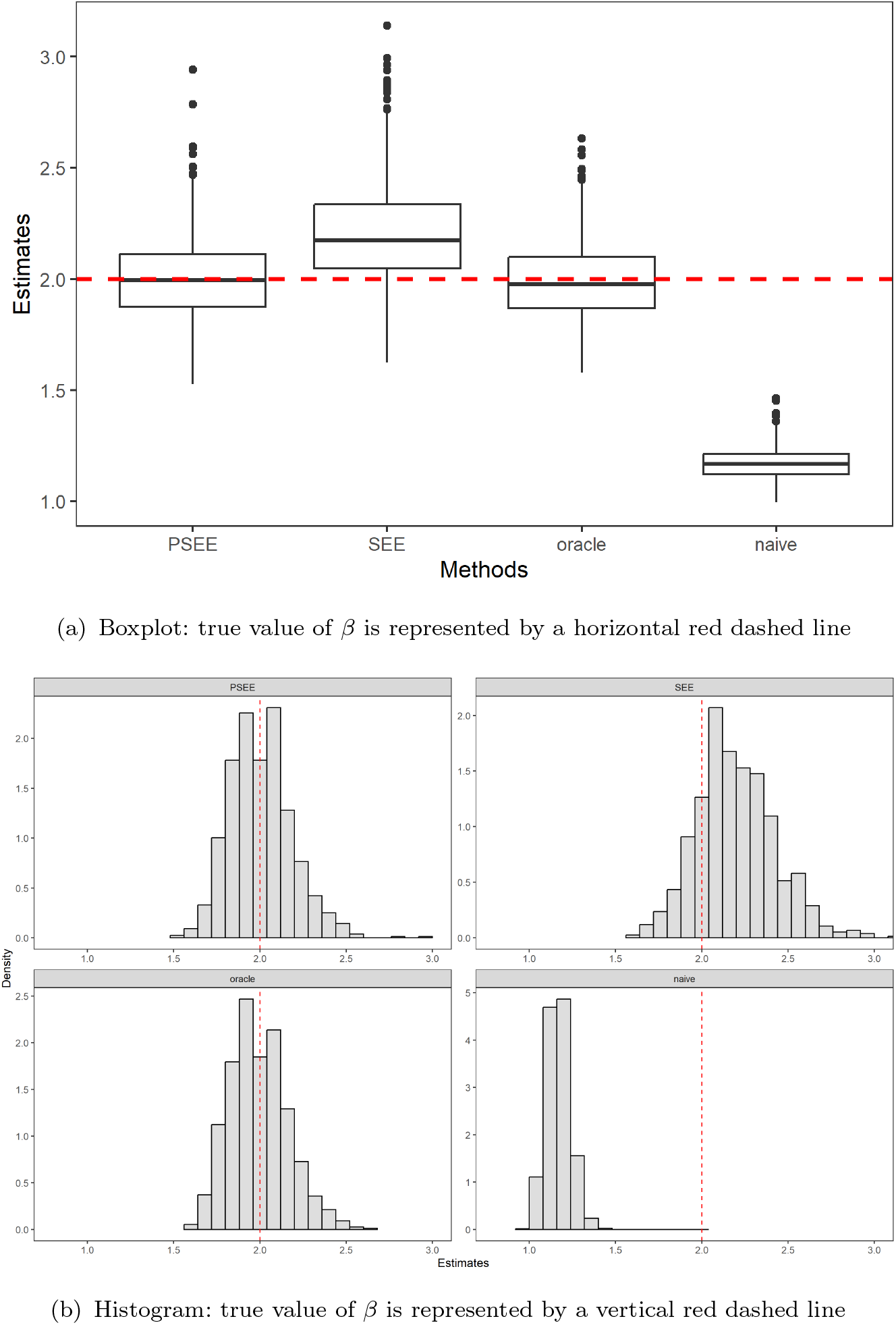
Boxplot and histogram of estimates of the causal effect *β, n* = 1000, *q* = 10, *U* ∼ *N* (0, 1), *c*_1_ = *c*_2_ = 0.5, 3 invalid IVs, 1000 replications.

**Figure A15:**
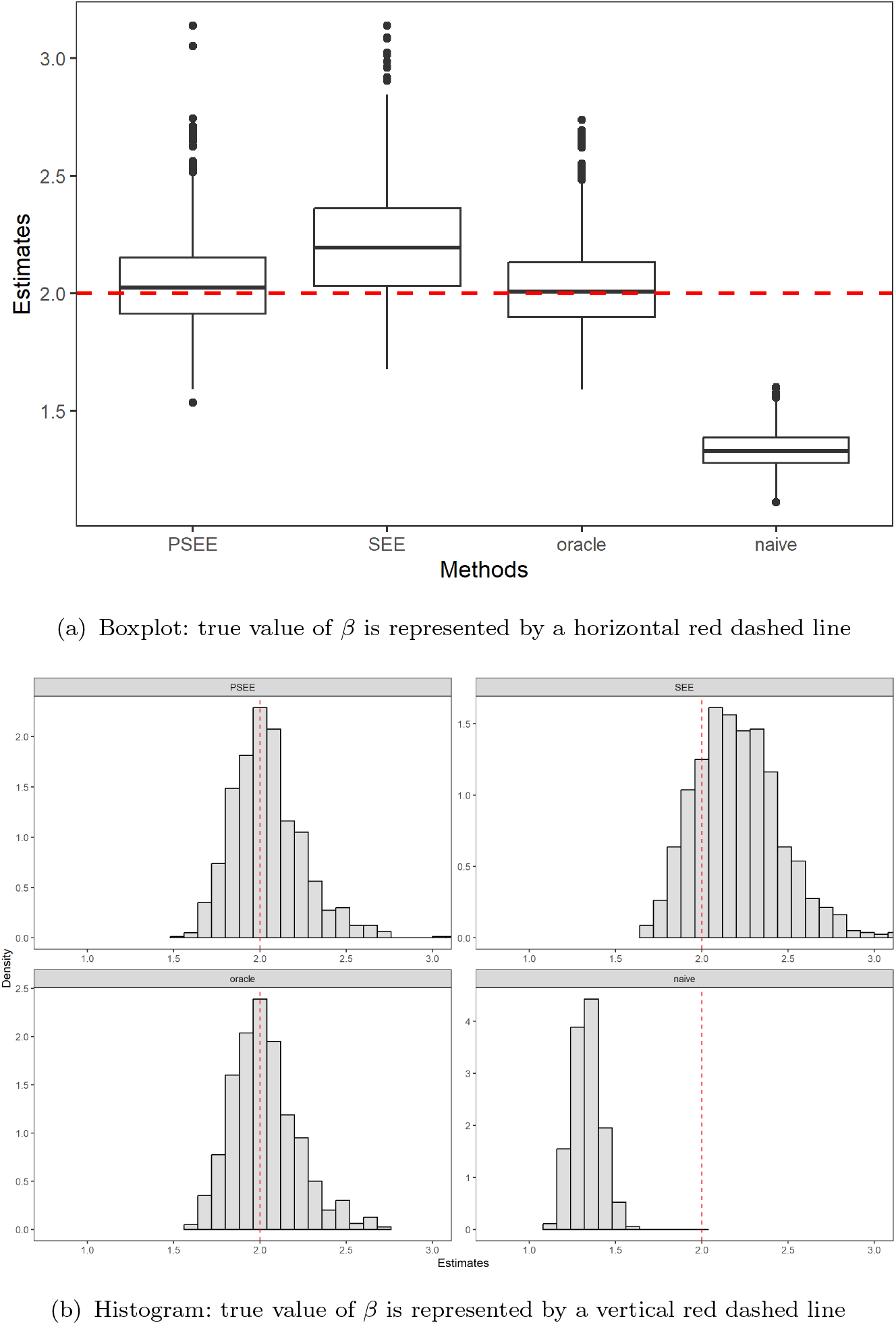
Boxplot and histogram of estimates of the causal effect *β, n* = 1000, *q* = 10, *U* ∼ *N* (0, 1), *c*_1_ = *c*_2_ = 0.5, 5 invalid IVs, 1000 replications.

**Figure A16:**
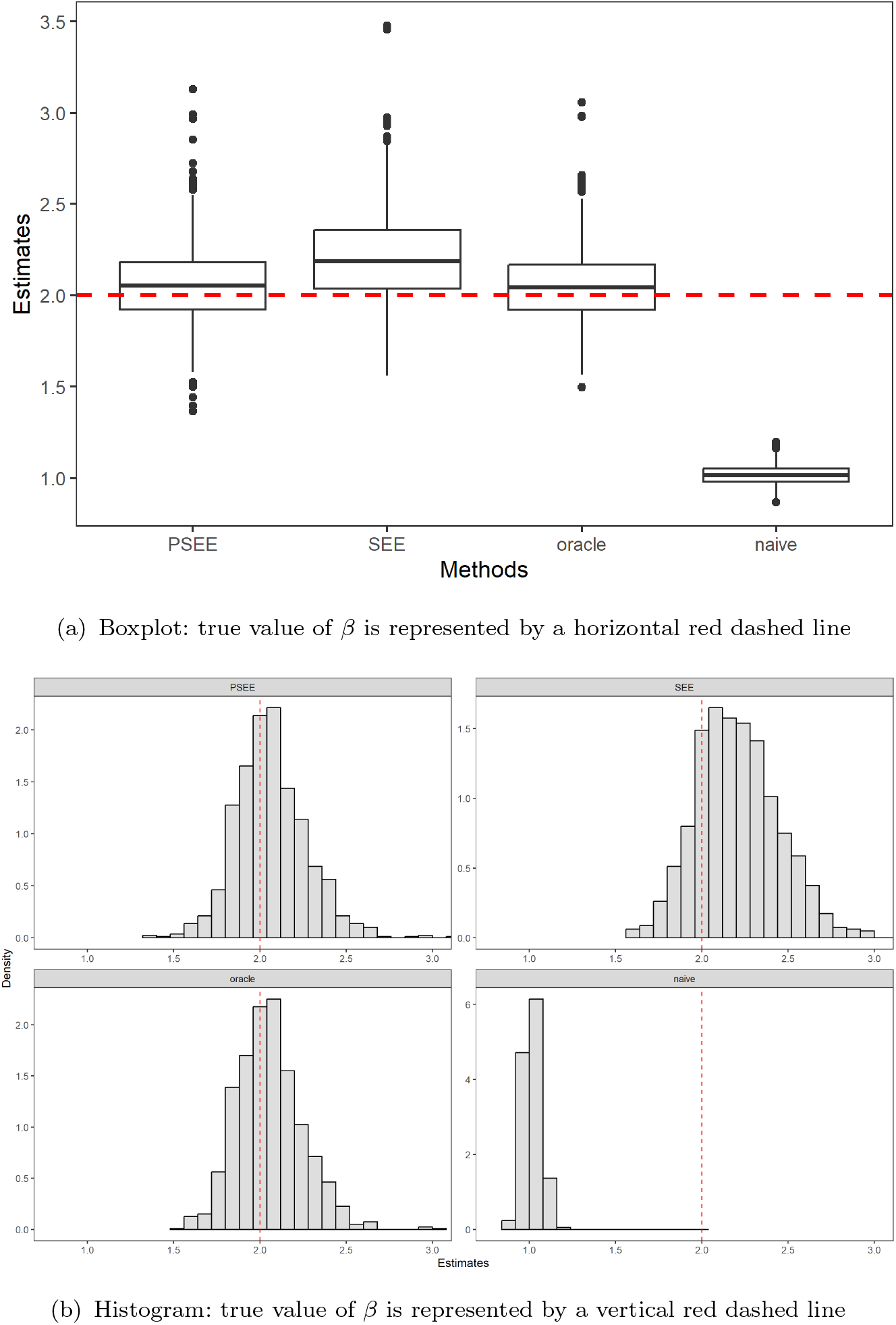
Boxplot and histogram of estimates of the causal effect *β, n* = 1000, *q* = 10, *U* ∼ *N* (0, 1), *c*1 = *c*2 = 0.5, 7 invalid IVs, 1000 replications.

*U ∼ t*(3) *and c*_1_ = *c*_2_ = 0.5.

**Table A2:**
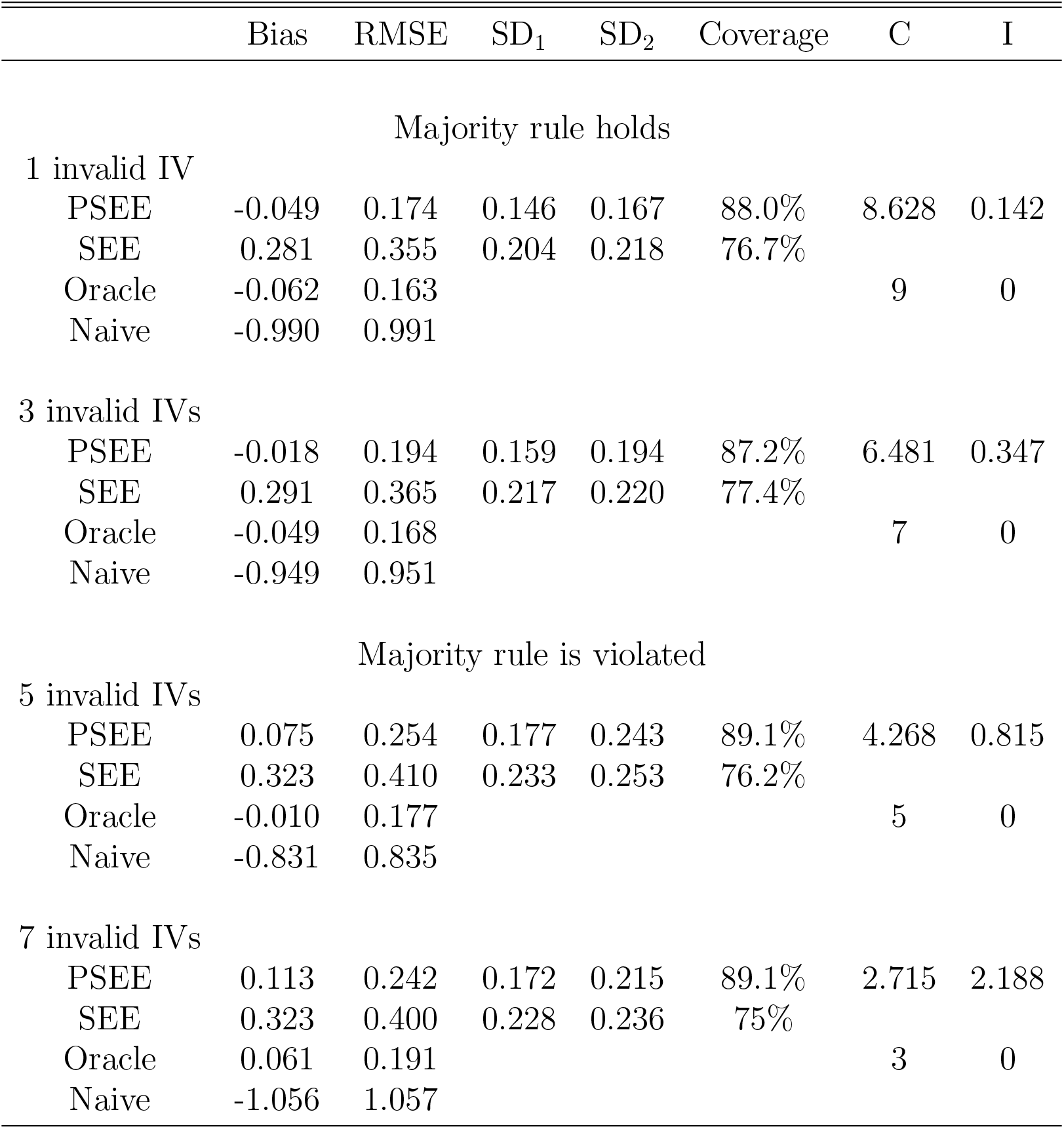
Estimation Results for *β* (*n* = 1000, *q* = 10, *U ∼ t*(3), *c*_1_ = *c*_2_ = 0.5): comparison of naive TSLS, oracle TSLS, PSEE, SEE. Working model for *U* is a discrete uniform distribution on the interval [*−* 0.5, 0.5] with mesh size *h*, here we take *h* = 0.5. SD_1_ and SD_2_ denote the sample standard deviation and the mean of the estimated standard deviation using the sandwich variance. C and I denote the average number of correct and incorrect zero estimates, respectively. True *β* equals 2.0.

| | Bias | RMSE | $SD_1$ | $SD_2$ | Coverage | C | I |
| --- | --- | --- | --- | --- | --- | --- | --- |
| Majority rule holds |  |  |  |  |  |  |  |
| 1 invalid IV |  |  |  |  |  |  |  |
| PSEE | -0.049 | 0.174 | 0.146 | 0.167 | 88.0% | 8.628 | 0.142 |
| SEE | 0.281 | 0.355 | 0.204 | 0.218 | 76.7% |  |  |
| Oracle | -0.062 | 0.163 |  |  |  | 9 | 0 |
| Naive | -0.990 | 0.991 |  |  |  |  |  |
| 3 invalid IVs |  |  |  |  |  |  |  |
| PSEE | -0.018 | 0.194 | 0.159 | 0.194 | 87.2% | 6.481 | 0.347 |
| SEE | 0.291 | 0.365 | 0.217 | 0.220 | 77.4% |  |  |
| Oracle | -0.049 | 0.168 |  |  |  | 7 | 0 |
| Naive | -0.949 | 0.951 |  |  |  |  |  |
| Majority rule is violated |  |  |  |  |  |  |  |
| 5 invalid IVs |  |  |  |  |  |  |  |
| PSEE | 0.075 | 0.254 | 0.177 | 0.243 | 89.1% | 4.268 | 0.815 |
| SEE | 0.323 | 0.410 | 0.233 | 0.253 | 76.2% |  |  |
| Oracle | -0.010 | 0.177 |  |  |  | 5 | 0 |
| Naive | -0.831 | 0.835 |  |  |  |  |  |
| 7 invalid IVs |  |  |  |  |  |  |  |
| PSEE | 0.113 | 0.242 | 0.172 | 0.215 | 89.1% | 2.715 | 2.188 |
| SEE | 0.323 | 0.400 | 0.228 | 0.236 | 75% |  |  |
| Oracle | 0.061 | 0.191 |  |  |  | 3 | 0 |
| Naive | -1.056 | 1.057 |  |  |  |  |  |

**Figure A17:**
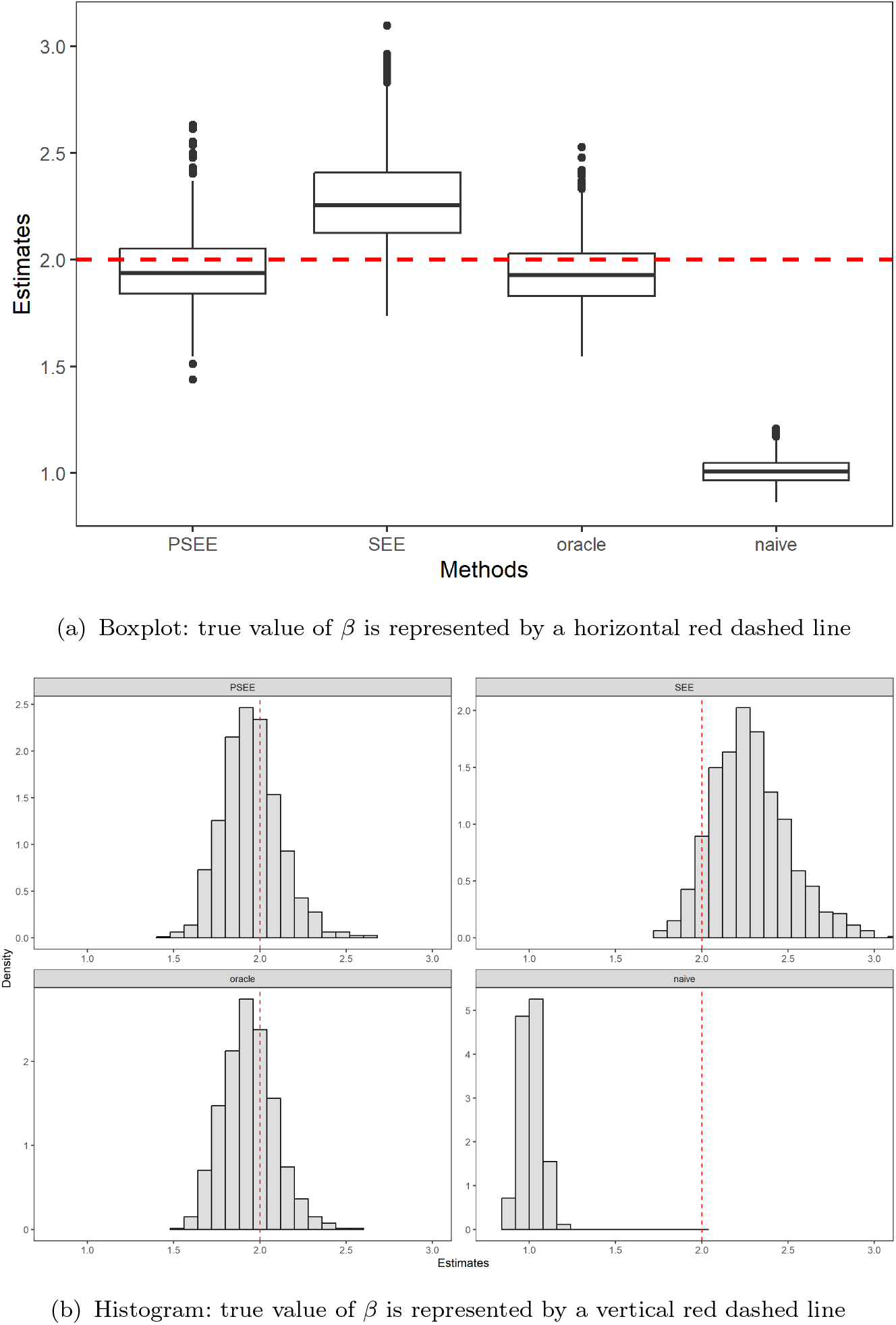
Boxplot and histogram of estimates of the causal effect *β, n* = 1000, *q* = 10, *U* ∼ *t* (3), *c*_1_ = *c*_2_ = 0.5, 1 invalid IV, 1000 replications.

**Figure A18:**
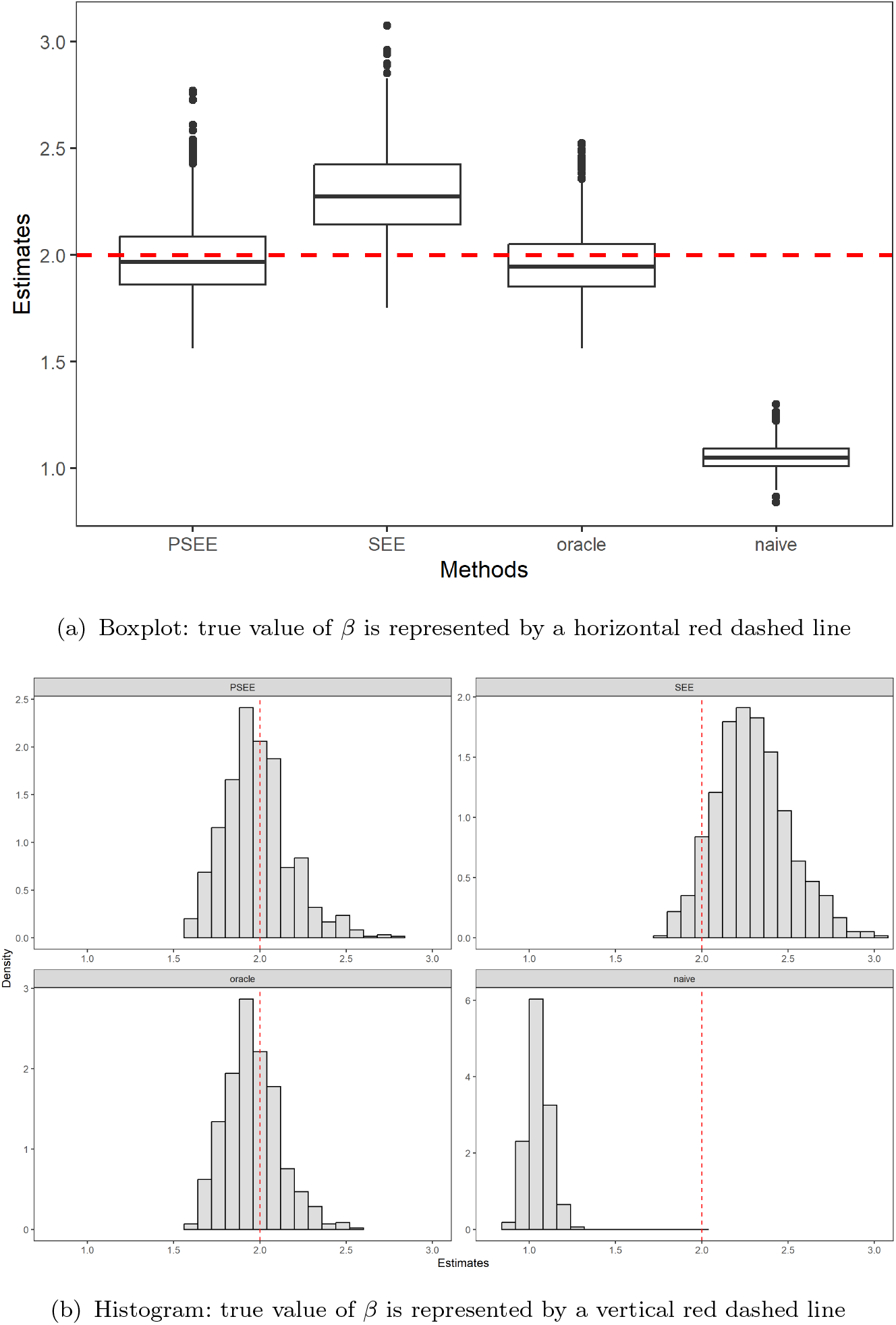
Boxplot and histogram of estimates of the causal effect *β, n* = 1000, *q* = 10, *U* ∼ *t* (3), *c*_1_ = *c*_2_ = 0.5, 3 invalid IVs, 1000 replications.

**Figure A19:**
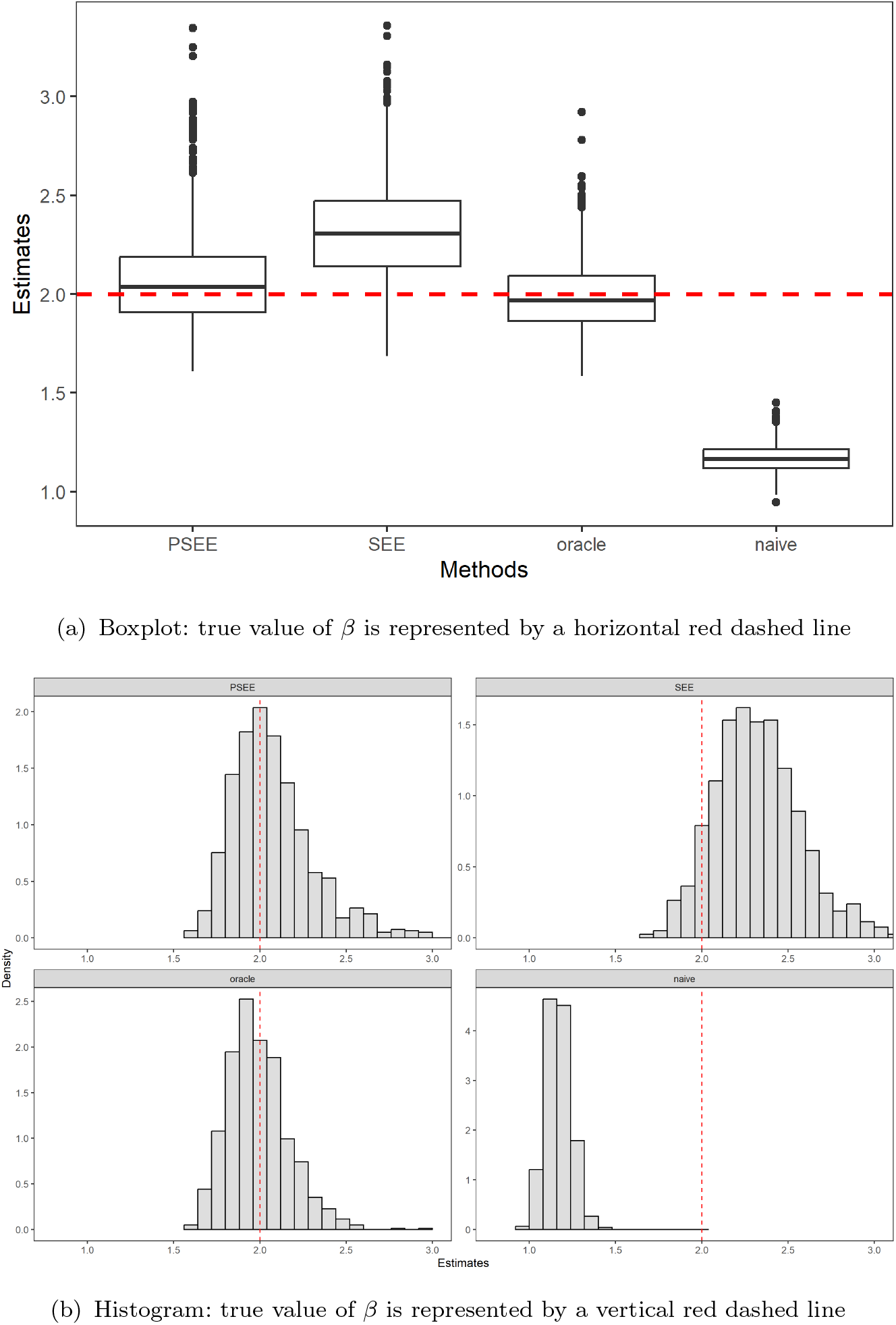
Boxplot and histogram of estimates of the causal effect *β, n* = 1000, *q* = 10, *U* ∼ *t*(3), *c*_1_ = *c*_2_ = 0.5, 5 invalid IVs, 1000 replications.

**Figure A20:**
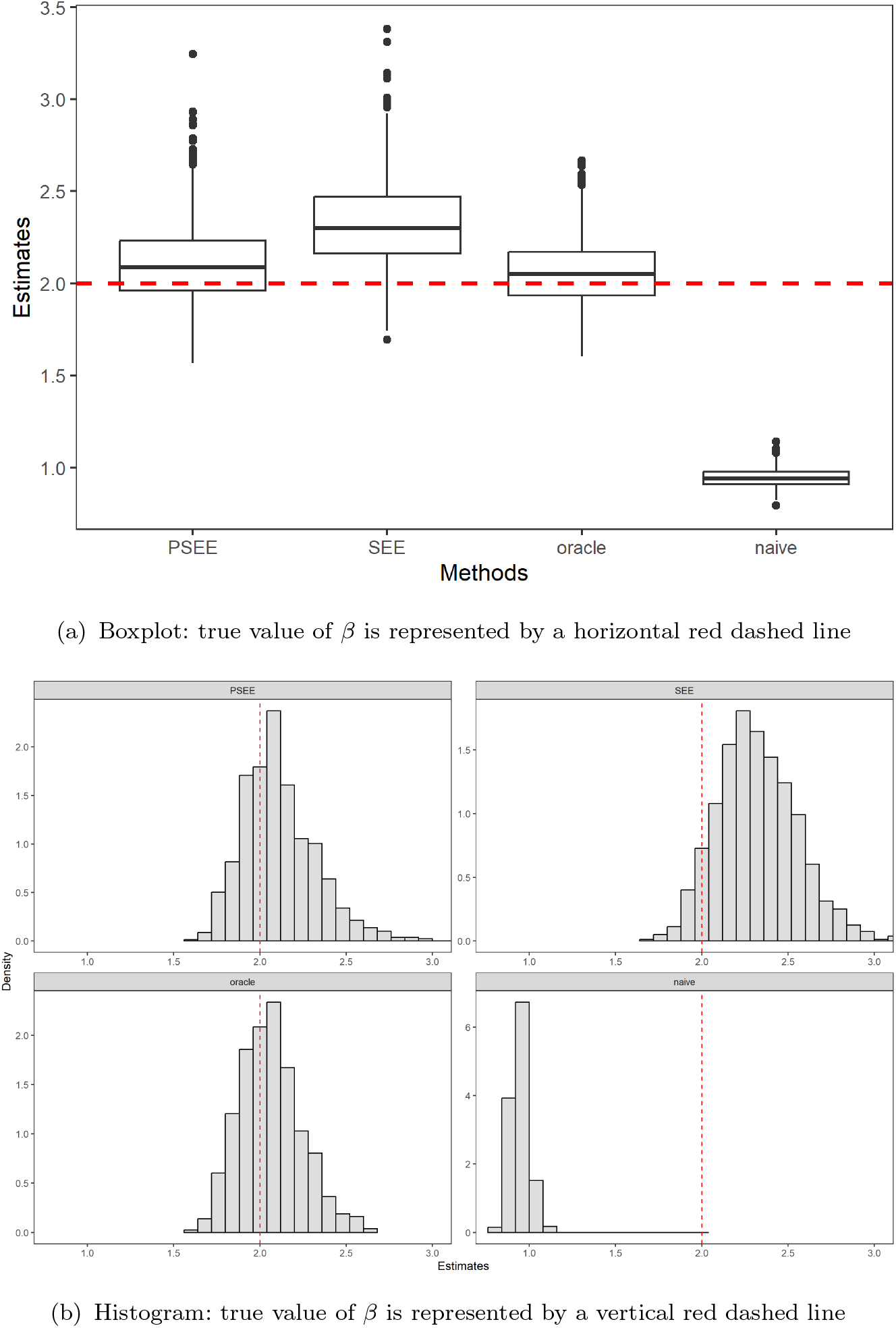
Boxplot and histogram of estimates of the causal effect *β, n* = 1000, *q* = 10, *U* ∼ *t*(3), *c*_1_ = *c*_2_ = 0.5, 7 invalid IVs, 1000 replications.

## Notes

### Competing Interest Statement

The authors have declared no competing interest.

### Author Declarations

Fudan University gave ethical approval for this work

